# Relationships of Alzheimer’s disease and apolipoprotein E genotypes with small RNA and protein cargo of brain tissue extracellular vesicles

**DOI:** 10.1101/2020.12.12.20247890

**Authors:** Yiyao Huang, Tom A. P. Driedonks, Lesley Cheng, Andrey Turchinovich, Harinda Rajapaksha, Tanina Arab, Bonita H. Powell, Olga Pletniková, Javier Redding, Juan C. Troncoso, Laura Vella, Lei Zheng, Andrew F. Hill, Vasiliki Mahairaki, Kenneth W. Witwer

**Affiliations:** Department of Molecular and Comparative Pathobiology, Johns Hopkins University School of Medicine, Baltimore, MD, USA; Department of Biochemistry and Genetics, La Trobe Institute for Molecular Science, La Trobe University, Bundoora, Australia; Molecular Epidemiology, German Cancer Research Center DKFZ, Heidelberg, Germany; SciBerg e.Kfm, Mannheim, Germany; Department of Pathology, Johns Hopkins University School of Medicine, Baltimore, MD, USA; Department of Pathology and Anatomical Sciences, Jacobs School of Medicine and Biomedical Sciences, University at Buffalo, Buffalo, NY, USA; Department of Neurology, Johns Hopkins University School of Medicine, Baltimore, MD, USA; The Florey Institute of Neuroscience and Mental Health, The University of Melbourne, Parkville, Australia; Department of Surgery, The University of Melbourne, The Royal Melbourne Hospital, Parkville, Australia; Department of Laboratory Medicine, Nanfang Hospital, Southern Medical University, Guangzhou, Guangdong, China; Richman Family Precision Medicine Center of Excellence in Alzheimer’s Disease, Johns Hopkins University School of Medicine, Baltimore, MD, USA

**Keywords:** Alzheimer’s disease, APOE, brain, extracellular vesicles, proteomics, RNA sequencing, exosomes, ectosomes, microvesicles

## Abstract

Alzheimer’s disease (AD) is a public health crisis that grows as populations age. Hallmarks of this neurodegenerative disease include aggregation of beta-amyloid peptides and hyperphosphorylated tau proteins in the brain. Variants of the APOE gene are the greatest known risk factors for sporadic AD. As emerging players in AD pathophysiology, extracellular vesicles (EVs) contain proteins, lipids, and RNAs and are involved in disposal of cellular toxins and intercellular communication. AD-related changes in the molecular composition of EVs may contribute to pathophysiology and lend insights into disease mechanisms. We recently adapted a method for separation of brain-derived EVs (bdEVs) from post-mortem tissues. Using this method, we isolated bdEVs from AD patients with different APOE genotypes and controls. bdEVs were counted, sized, and subjected to parallel small RNA sequencing, proteomic analysis. Although overall bdEV concentration was not affected by AD, we observed a shift towards smaller particles in AD. Also, numerous bdEV-associated RNAs (including miRNAs and tRNAs) and proteins were found to be correlated with AD pathology and APOE genotype. Some of the identified entities have been implicated previously in important AD-related pathways, including amyloid processing, neurodegeneration, and metabolic functions, etc. Prominently, AD hallmark Tau and Tau phosphorylated at threonine 231 (phosTau) were significantly increased in AD bdEVs, indicating the involvement of bdEVs in spread of Tau pathology. These findings provide further evidence that bdEVs and their molecular cargo modulate development and progression of AD.

## INTRODUCTION

Alzheimer’s disease (AD, [194]) is a public health crisis [5, 141] that continues to grow as the population ages, demanding new insights into pathophysiology. Mechanisms of pathogenesis are manifold and incompletely understood. Synaptic connections are lost through neurodegeneration. Extracellular deposition of β-amyloid (Aβ—fragments of the amyloid precursor protein, APP) occurs in the form of neuritic plaques (NPs), while intracellular aggregation of hyperphosphorylated tau produces neurofibrillary tangles (NFTs) [114, 121, 154, 186, 187]. Similar to misfolded prions, pathogenic Aβ and tau can template misfolding and protein modifications at recipient sites [3, 157, 179]. Wide distribution and high density of NPs and NFTs correspond with greater cognitive impairment. Other hallmarks of AD include vascular amyloidosis, gliosis, and blood-brain barrier disruption [173].

Numerous genetic risk factors for AD have been identified. Mutations in genes such as presenilin 1 (PSEN1) and APP contribute to early-onset, familial AD (FAD). In sporadic AD, variants of the apolipoprotein E (APOE) gene (APOE) are the greatest known genetic risk factor [105, 147, 158, 163]. A central player in lipid homeostasis, APOE assists in intercellular lipid transfer, binding both cholesterol and low-density lipoprotein receptors in addition to binding Aβ. Three major APOE isoforms exist, encoded by alleles ε2, ε3, and ε4, and produce proteins that differ by only 1-2 amino acids but have different binding partner interactions. Whereas APOE2 is protective against AD relative to APOE3, APOE4 is associated with increased risk, particularly in women and certain ethnic populations [54]. In Caucasians, the risk of late-onset AD is higher for APOE4 homozygosity (12-fold) and heterozygosity (2-3-fold), with some indication of greater risk for early-onset disease [143]. A majority of AD patients in the US have at least one ε4 allele. The critical role of APOE in central nervous system (CNS) lipid metabolism is not limited to AD. The ε4 allele has been associated with increased cholesterol, ceramide, and sphingomyelin in brains of HIV dementia patients [39], a difference not due to inflammation (activated microglia) or astrogliosis. Furthermore, interactions of biological sex (AD disproportionately affects women [80]) and APOE4 have been noted in AD [54, 127, 168] and AD models [24, 137].

Extracellular vesicles (EVs) comprise a diversity of lipid bilayer membrane-delimited particles that dispose cellular toxins and mediate intercellular communication [97, 162, 181]. Roles for EVs have recently been recognized in AD pathophysiology [45, 170, 172]. EVs participate in neurodegeneration in AD, Parkinson’s, and prion diseases in part by spreading misfolded proteins [50, 51, 130, 138, 145]. In AD, Aβ and tau have been found in or on EVs in AD models and patients [11, 50, 55, 69, 130, 134, 139, 145, 146, 159]. EVs are also found “at the scene of the crime”: in amyloid plaques [139, 140]. EV cargo betrays AD and mild cognitive impairment (MCI) [55, 61, 66, 89, 93, 103, 159]. Furthermore, some EVs contribute to amyloid clearance by glia [6, 188, 189], suggesting that EVs from healthy cells block AD pathology. However, context matters: in certain models, reducing EV release may diminish pathology [43, 44]. Differential expression of various microRNAs was observed in brain tissue [37, 99, 177] and serum of AD patients [29], suggesting that the RNA cargo of EVs may contribute to AD pathogenesis.

The study of EVs and APOE, like the study of EVs in AD in general, is just beginning [1, 124, 129, 169], but the rapidly growing literature shows that EVs and APOE contribute to AD via multiple mechanisms. At the same time, APOE variants act in lipid homeostasis and trafficking, processes that also govern EV release and uptake. APOE+ EVs have recently been characterized in cerebrospinal fluid (CSF) and cell culture, with changes in fluids of AD and MCI patients [185]. In the pigment cell amyloid model system [178], APOE was associated with EVs in the multivesicular body, where it nucleated formation of cytotoxic amyloid aggregates [123, 169] before release from the cell. APOE is prominently sorted into EVs, particularly from neurons and astrocytes [124]. APOE shuttling may increase upon exposure to Aβ peptides [124]. Consistent with an influence on endosome-origin EV biogenesis, APOE is a well-known determinant of transmission of certain enveloped viruses, a role exerted in the cell of origin, not the recipient cell [192]. Studies in the periphery have touched on the role of EVs in the context of APOE and atherosclerosis (e.g., [62, 104, 133]), but without finding direct connections.

In this study, we separated and characterized EVs from human AD and control brain tissue. Most studies of EVs in AD to date focus on EV functions or EV-associated biomarkers in biofluids or cell culture supernatant. However, EVs are most likely to act locally, in the tissue of origin. We therefore obtained brain tissue of AD cases with different APOE genotypes as well as control brain tissue. We used our modification of a rigorous method for separation of tissue EVs [85, 171] to obtain and characterize bdEVs in accordance with the recommendations of the Minimal Information for Studies of EVs (MISEV) and related initiatives of the International Society for Extracellular Vesicles (ISEV) [42, 108, 162, 180]. RNA and protein profiling revealed multiple molecular entities associated with Alzheimer’s disease and, to a lesser extent, with APOE genotype in AD.

## METHODS

### Tissue collection, processing, and approvals

Human brain tissues were obtained from Johns Hopkins Alzheimer’s Disease Research Center. All collections were approved by the Johns Hopkins University Institutional Review Board. Following external examination and weighing of the autopsy brain, the right cerebral hemisphere was cut in coronal slabs and frozen on prechilled metal plates and stored at –80°C. A mixture of temporal cortex and inferior parietal cortex tissues from two cohorts of brains were used in this study. The first discovery cohort including 23 Alzheimer’s disease (AD) patients and 7 non-AD controls (Table S1), while the second validation cohort including 21 Alzheimer’s disease (AD) patients and 10 non-AD controls (Table S2).. AD patients were diagnosed according to Braak [21] and CERAD [115, 117] criteria. For APOE genotyping, genomic DNA was extracted from tissue using the DNeasy kit (Qiagen). Genotyping was performed using standard procedures [83].

### Separation of extracellular vesicles from brain tissue

EVs were separated from brain tissues using our published protocol [85]. Before extraction, a small (∼50 mg) piece of tissue was stored at −80°C for later protein and RNA extraction. The remaining frozen tissue was weighed and briefly sliced on dry ice and then incubated in 75 U/ml collagenase type 3 (Worthington #CLS-3, S8P18814) in Hibernate-E solution for 20 min at 37°C. PhosSTOP and Complete Protease Inhibitor (SigmaAldrich PS/PI 4906837001/11697498001) solution were added to stop digestion. The dissociated tissue was spun at 300 × g for 10 min at 4°C. Supernatant was transferred to a fresh tube and spun at 2000 × g for 15 min at 4°C. Supernatant was filtered through a 0.22 μm filter (Millipore Sigma, SLGS033SS) to deplete tissue debris and spun at 10,000 × g for 30 min at 4°C (Thermo Fisher swinging-bucket rotor model AH-650, k-factor 53, acceleration and deceleration settings of 9, using Ultra-Clear tubes with 5 ml capacity). The pellet was resuspended in 100 μl PBS. This resuspension is termed the “10,000 × g pellet” (10K). The 10,000 × g supernatant was concentrated by 100 kilodalton (kDa) MWCO protein concentrator (Thermo Fisher 88524) from 5 ml to 0.5 ml and loaded onto a size-exclusion chromatography column (qEV original, IZON Science SP1-USD, Christchurch, New Zealand) and eluted by PBS. 0.5 ml fractions were collected. The first 3 ml (Fractions 1-6) of the eluate were discarded as the void volume and subsequent 0.5 mL fractions were collected. For the purposes of this study, a total of 2 ml eluate (Fractions 7-10) were pooled and ultracentrifuged for 70 min at 110,000 × g (average) at 4°C (swinging-bucket rotor model TH-641, Thermo Fisher, k factor 114 at max speed, acceleration and deceleration settings of 9, using thinwall polypropylene tubes with 13.2 ml capacity). Supernatant was removed, and the pellet was resuspended in 100 μl PBS as the purified EV fraction. Fractions were stored at −80°C.

### Brain homogenate preparation for protein and RNA

For protein extraction, brain homogenates (BH) were prepared by grinding tissue in cold PBS containing PI/PS with a handheld homogenizer (Kontes Pellet Pestle Motor) for 10 sec. RIPA lysis buffer (Cell Signaling Technology 9806) was added and the mixture was sonicated using an ultrasonic ice bath at 20 kHz for 4 × 20 sec, with a 10 sec interval between each sonication. Homogenate was rotated at 4°C for 2 h and spun 15 min at 14,000 × g at 4°C. Supernatant was transferred to tubes and stored at −80°C.

For RNA extraction, Trizol (Thermo Fisher 15596018) was added to frozen brain tissue and homogenized with Lysing Matrix D beads (MP Biomedicals 116913100) using a benchtop homogenizer at speed setting of 4.0 m/s for 20 sec (FastPrep-24, MP Biomedicals). After homogenization, the supernatant was collected and subjected to RNA extraction by miRNeasy Mini Kit (Qiagen 217004) according to the manufacturer’s instructions.

### Nano-flow analysis

EV concentration and size distribution were estimated using the nanoFCM flow nanoAnalyzer (NanoFCM Co.) per manufacturer’s instructions. The instrument was calibrated for concentration and size using 200 nm polystyrene beads and a silica nanosphere cocktail (provided by NanoFCM as pre-mixed silica beads with diameters of 68, 91, 113, and 151 nm), respectively. EV and 10K preparations were resuspended in 50 μl of PBS, diluted as needed (typically 1:200 dilution for EV and 1:50 dilution for 10K fractions), and loaded and recorded for 1 minute. Using a calibration curve, the flow rate and side scatter intensity were converted into particle numbers.

### RNA extraction and quality control

RNA was extracted by miRNeasy Mini Kit (Qiagen 217004) according to the manufacturer’s instructions. The small RNA profiles of samples were analyzed using capillary electrophoresis by RNA 6000 Pico Kit (Agilent Technologies 5067-1513) on a Fragment Analyzer (Advanced Analytical). Total RNA and small RNA from BH were analyzed using capillary electrophoresis by RNA 6000 Nano Kit (Agilent Technologies 5067-1511) and RNA 6000 Pico Kit.

### Small RNA sequencing

EV RNA was concentrated to 6 µl using the Savant SpeedVac Vacuum concentrator. Small RNA libraries were constructed from 50 ng of RNA extracted from brain homogenate or 5 µl of RNA from bdEVs using the Ion Total RNA-Seq Kit V2 (Life Technologies 4475936). Barcodes were attached using the Ion Xpress™ RNA-Seq Barcode 1-16 Kit (Life Technologies 4471250) according to the manufacturer’s protocol and as previously published [30]. The yield and size distribution of the small RNA libraries (96nt to 250nt) were assessed using the Agilent 2100 Bioanalyzer™ instrument with DNA 1000 chip (Agilent Technologies 5067-1504). Multiplexed libraries were equally pooled to 45 pM and prepared for deep sequencing using the Ion Chef system (Life Technologies 4484177) and sequenced on the Ion Torrent S5™ using Ion™ 540 chips (Life Technologies A27765).

### RNA sequencing data analysis

Original BAM files were converted into FASTQ format using picard tools (SamToFastq command). Reads shorter than 15 nt were removed from the raw FASTQ data using cutadapt software v1.18. The size-selected reads were aligned to human reference transcriptomes using bowtie software (1 mismatch tolerance) in a sequential manner. Specifically, reads were first mapped to rRNA, tRNA, RN7S, snRNA, snoRNA, scaRNA, VT-RNA, Y-RNA as well as the mitochondrial genome. All reads which did not map to the above RNA species were aligned to human miRNA references (miRBase 22 release). The remaining reads were further aligned to protein-coding mRNAs and long non-coding RNA (lncRNA) references (GENCODE Release 29). The numbers of reads mapped to each RNA type were extracted using eXpress software based on a previous publication [142]. To compare the RNA biotype distribution, reads of different RNA biotypes were normalized as reads per million total reads (RPM). Differential gene expression was quantified using R/Bioconductor packages DESeq or DESeq2 as described [7, 109].

### Mass spectrometry

Samples were resuspended in 1 X RIPA buffer (20mM Tris-HCl pH7.5, 150mM NaCl, 1mM Na2EDTA, 1mM EGTA, 1% NP-40, 1% SDS, 2.5mM sodium pyrophosphate, 1mM β-glycerophosphate, 1mM Na3VO4, 1μg/ml leupeptin) and protease inhibitors and incubated on ice for 5 min. The samples were sonicated for 15 min in an ice water bath before centrifugation at 14,000 × g at 4°C for 10 min. The supernatant was collected and assessed for protein concentration using the micro BCA assay (Thermo Fisher Scientific 23235). 3 μg of brain homogenate and 1.5 µg of 10K pellet and EV samples were buffer-exchanged prior to mass spectrometry to remove detergent. Proteins were resuspended in 8M Urea, 50 mM Tris pH=8.3. 1 µL of TCEP (tris [2-carboxyethyl] phosphine hydrochloride, 200 mM solution in water) was then added to the samples and incubated for 4 hours at 21°C in a ThermoMixer (Eppendorf AG). 4 µL of 1M IAA (iodoacetamide in water) was then added and samples were incubated in the dark at 21°C. 800 µL of 50 mM Tris (pH 8.3) and 1 μg trypsin were then added to samples prior to overnight incubation at 37°C. 10 µL of 10% trifluoroacetic acid (TFA) was added to each sample to acidify. Samples were cleaned using stage-tips preparations using 3 plugs of Empore polystyrenedivinylbenzene (SBD-XC) copolymer disks (Sigma Aldrich, MO, USA) for solid phase extraction. Peptides were reconstituted in 0.1% formic acid and 2% acetonitrile and loaded onto a trap column (C18 PepMap 100 μm i.d. × 2 cm trapping column, Thermo Fisher Scientific) at 5 µL/min for 6 min using a Thermo Scientific UltiMate 3000 RSLCnano system and washed for 6 min before switching the precolumn in line with the analytical column (BEH C18, 1.7 μm, 130 Å and 75 μm ID × 25 cm, Waters). Separation of peptides was performed at 45°C, 250 nL/min using a linear ACN gradient of buffer A (water with 0.1% formic acid, 2% ACN) and buffer B (water with 0.1% formic acid, 80% ACN), starting from 2% buffer B to 13% B in 6 min and then to 33% B over 70 min followed by 50% B at 80 min. The gradient was then increased from 50% B to 95% B for 5 min and maintained at 95% B for 1 min. The column was then equilibrated for 4 min in water with 0.1% formic acid, 2% ACN. Data were collected on a Q Exactive HF (Thermo Fisher Scientific) in Data Dependent Acquisition mode using m/z 350–1500 as MS scan range at 60 000 resolution. HCD MS/MS spectra were collected for the 7 most intense ions per MS scan at 60 000 resolution with a normalized collision energy of 28% and an isolation window of 1.4 m/z. Dynamic exclusion parameters were set as follows: exclude isotope on, duration 30 s, and peptide match preferred. Other instrument parameters for the Orbitrap were MS maximum injection time 30 ms with AGC target 3 × 10^6^, MSMS for a maximum injection time of 110 ms with AGT target of 1 × 10^5^.

### Proteomics data analysis

Human protein sequences (last modified date: 16 May 2019) were downloaded from the Uniprot database and used as the database for the search engine. Common Repository of Adventitious Proteins (CRAP) was used as the potential lab contaminant database. Protein identification was performed using the proteomics search engine Andromeda built in to Maxquant V 1.16.0. Trypsin with a maximum of two missed cleavages was used as the cleavage enzyme. Carbamidomethyl of cysteine was set as fixed modification and oxidation of methionine was set as variable modification. The false discovery rate (FDR) was set to 1%. The Label Free quantification was done with match between runs using a match window of 0.7 min. Large label free quantification (LFQ) ratios were stabilized to reduce the sensitivity for outliers. For human datasets, data scaling was done using the cyclic loess method, and scaled data were visualized with a PCA plot. For differential abundance analysis, nested factorial design was set up for the analysis, where each subtype of the disease was nested within the main disease category and contrasts for the main categories were computed by averaging the subtypes.

The protein interaction and cluster protein function prediction was done by Protein-Protein Interaction Networks Functional Enrichment Analysis (STRING) [161]. Kyoto Encyclopedia of Genes and Genomes (KEGG) [92] was used to enrich pathway involvement of identified proteins. Statistical significance of enrichment was determined by the tools mentioned above. Only nominally significant categories (false discovery rate (FDR) < 0.05) were included for analysis.

### Electrochemiluminescence-linked (ECL) immunoassay for total tau and phosphorylated tau detection

Total tau and phosphorylated tau at threonine 231 (phosTau) were measured in BH, 10K, and EVs using an ECL-immunoassay (Meso Scale Discovery K15121D) according to the manufacturer’s instructions. In brief, 10K and EV samples were diluted 1:10 while BH was diluted 1:100 in 2% block buffer (2% BSA in tris buffer) containing 0.5% triton X-100. Samples were incubated for one hour on the plate. After washing the plate, the SULFO-TAG Anti-Total Tau Antibody was added and incubated with the plate for one hour. After washing, MSD Read Buffer was added, and the plate was read immediately with a Quick plus SQ 120 MM instrument. Data analysis was done on MSD DISCOVERY WORKBENCH software version 2.0.

### Statistical analysis

Statistical significance of particle count, particle: protein ratio, size distribution, RNA biotype differences, and tau and phosTau level differences between AD and control groups and between either of two APOE genotype groups were determined by two-tailed Welch’s t-test. Receiver operating characteristic (ROC) analyses of Tau and phosTau between AD and control were generated in GraphPad Prism 8.1 using the method of Wilson and Brown.

### Data availability

Nucleic acid sequencing data have been deposited with the Gene Expression Omnibus, accession GSE159541. Proteomics data files are available on request. We have submitted all relevant details of our experiments to the EV-TRACK knowledgebase (EV-TRACK ID: EV200126) [42].

## RESULTS

### Separation of EVs from AD and control brain tissue

Following the protocol illustrated in Fig. 1a, we separated bdEVs from brain tissue of controls (n=7) and individuals with AD (n=23) and different APOE genotypes (Table 1, Table S1, S2). A small amount (∼50 mg) of each tissue was set aside to produce brain homogenate (BH) to assess protein and RNA profiles of the source material. After enzymatic digestion and initial filtering of the remaining tissue, 10,000 x g ultracentrifuged pellets were collected and termed “10K” as an intermediate product of EV separation. The 10K supernatant was then separated by size exclusion chromatography (SEC) [85] and concentrated into a more pure EV preparation. Fractions were processed for characterization, including proteomics and small RNA profiling.

**Fig. 1.**
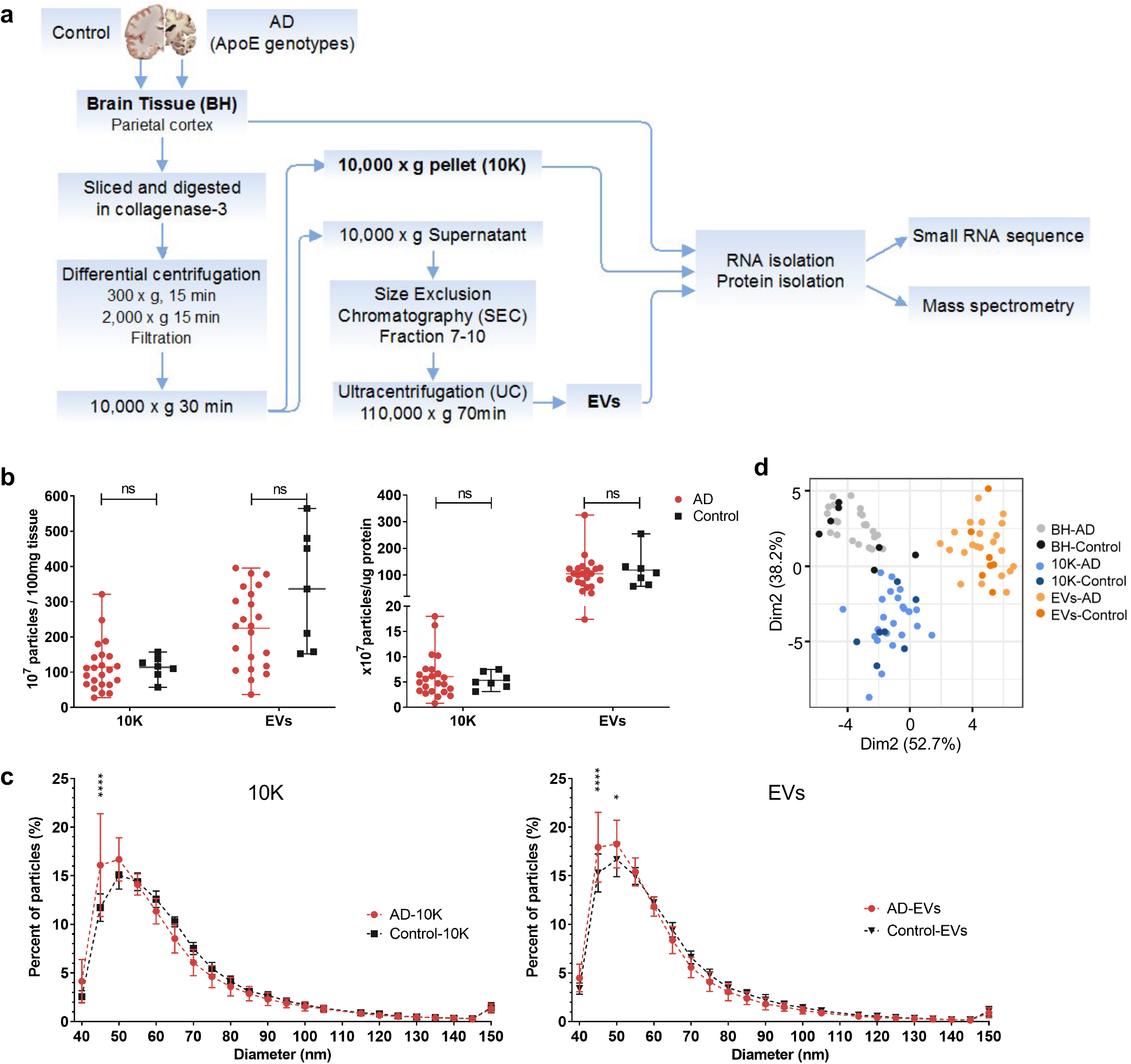
Alzheimer’s disease (AD) and control brain tissue-derived EV (bdEV) enrichment and characterization. (a) Workflow for bdEV enrichment, small RNA sequencing and proteomics. Following digestion, centrifugation, and filtration steps, 10,000 x g pellets from AD and control brain tissue (BH) (as indicated in Table 1) were collected and defined as the 10K fraction. Size-exclusion chromatography (SEC) was applied to 10,000 x g supernatants to enrich EVs. RNA and proteins from BH, 10K, and EVs were then isolated and subjected to small RNA sequencing and mass spectrometry. (b) Left: Particle concentrations of 10K and EV fractions of AD and control samples were measured by NFCM. Particle concentration for each group was normalized by tissue mass (per 100 mg). Right: Ratio of particles to protein (particles/µg). Protein concentrations of 10K and EV fractions were measured by BCA protein assay. Data are mean +/- SD. ns: no significant difference (p > 0.05) between AD and control by two-tailed Welch’s t-test. (c) Size distributions of 10K and EV fractions (AD and control) were measured by NFCM and calculated as particles in a specific size bin versus total detected particles in each sample (percentage). Data are presented as mean +/- SD. *p ≤ 0.05, ****p ≤ 0.0001 by two-tailed Welch’s t-test. (d) Principal component analysis (PCA) based on extracellular vesicle (EV) marker expression per proteomics analysis. EV markers used for PCA: CD81, CD9, FLOT1, FLOT2, RAB1A, RAB7A, TUBA1B, TUBB4B, ANXA2, ANXA5, ANXA6, ACTN1, GAPDH. Data are presented as mean +/- SD.

**Table 1.** Donor characteristics: Cohorts 1 and 2.

| Cohort 1 |  |  |  |
| --- | --- | --- | --- |
| For profile | AD (n=23) | Control (n=7) | p |
| Age, mean | 77.4 ± 12.91 | 82.4 ± 12.47 | 0.3749 |
| Sex (male, female) | 9M, 13F | 4M, 3F |  |
| PMI, mean | 10.8 ± 5.63 | 8.0 ± 3.60 | 0.136 |
| Cohort 2 |  |  |  |
| For Tau and phosTau<sup>a</sup><br>detection | AD (n=21) | Control (n=10) | p |
| Age, mean | 78.8 ± 12.45 | 78.3 ± 12.60 | 0.9248 |
| Sex (male, female) | 8M, 13F | 7M, 3F |  |
| PMI, mean | 10.7 ± 5.43 | 9.2 ± 5.22 | 0.4657 |

<sup>a</sup> phosphorylated Tau

### Basic EV characterization

Particle concentration per 100 mg tissue input and particle size distribution were determined by nano-flow cytometry measurement (NFCM). No significant particle yield difference was detected between AD and control brain-derived 10K and EV fractions (Fig. 1b left). Particle:protein ratio was also calculated to evaluate EV purity. This ratio was similar between the AD and control groups (Fig. 1b right). However, the EV fraction had a higher particle:protein ratio compared with 10K, as expected and consistent with greater protein contamination of the 10K pellet (Fig. 1b right). Fractions from AD and control groups also had similar size distributions. However, more small particles in the approximately 45-50 nm diameter range were observed in AD samples compared with controls (Fig. 1c). No significant particle count differences or size distribution shifts were observed for samples with different APOE genotypes (ε2/3, ε3/3, ε3/4/, and ε4/4; Fig. S1a-b). We examined the expression levels of 13 proteins that are commonly reported to be associated with EVs (CD81, CD9, FLOT1, FLOT2, RAB1A, RAB7A, TUBA1B, TUBB4B, ANXA2, ANXA5, ANXA6, ACTN1, GAPDH). These proteins were not found to be significantly differentially abundant between AD and controls. Principal component analysis (PCA) also showed different EV marker expression patterns between BH, 10K, and EV fractions, but not between AD and controls (Fig. 1d).

### Assessment of RNA biotype distribution

Small RNA sequencing of BH, 10K, and EV fractions yielded 3.2M (± 1M), 2.2M (± 0.9M), and 0.75M (± 0.36M) reads, respectively (M = million, 1 x 10^6). After adapter clipping and removing reads shorter than 15 nt, 66% (± 3.9%) of BH, 63% (±10.6%) of 10K, and 31% (± 8.5%) of EV reads mapped to the human genome (hg38). Reads mapped to various RNA biotypes were normalized to reads per million mapped reads (RPM) (Fig. S2a-c). Overall, no major differences in biotype distribution were observed between AD and control conditions, although AD EVs showed a slight decrease in mitochondrial RNA (mtRNA) (Fig. S2c). We then determined enrichment of ncRNA biotypes in EVs relative to tissues and assessed whether the enrichment pattern differed between AD and control brains. To do this, we normalized the RPM of each ncRNA biotype to its average RPM in brain tissues (Fig. S2d). Consistent with earlier reports [46, 85, 125, 165], miRNA and snoRNA were relatively underrepresented in EVs compared with brain homogenates. The same was observed for mtRNA and, to a lesser extent, for snRNA. Y-RNA was equally abundant in BH, 10K, and EVs. tRNA was enriched in 10K and was equally abundant in EVs and BH. RN7S (SRP-RNA) was significantly enriched in the purified EV fraction of AD samples, but not of controls. Vault RNAs were strongly enriched in EVs, although this may have been due to co-purification of extracellular Vault particles with EVs, as reported earlier [88]. Interestingly, both protein-coding (mRNAs) and long non-coding RNAs (lncRNAs) were enriched in EVs compared with BH and 10K, and these reads mapped to thousands of different genes. However, because of our size selection and library preparation methods, these reads likely represent the RNA degradome and not full-length RNAs.

### EVs are associated with differentially expressed miRNAs related to AD pathology

Examining individual miRNAs, 26 miRNAs (10K) and 16 miRNAs (EVs) differed significantly (FDR < 0.05) between AD and controls (Fig. 2a). Of these, two miRNAs in 10K and nine miRNAs in EVs differed by more than two-fold (Fig. S3a-b). Table 2 provides a summary of these dysregulated miRNAs, indicating up/downregulation and involvement in AD as reported in previous studies, since up/downregulation of some EV-associated miRNAs was consistent with reports of brain tissue expression [37, 99, 177]. By APOE genotype, focusing on the presumed highest-risk ε4/4 and the lowest-risk e2/3 carriers in our study, nine miRNAs in 10K differed more than two-fold (with p value < 0.05), while only two miRNAs (miR-379-5p and miR-199a-5p) differed in EVs (Fig. 2b, S4a-b, and Table 3). However, none of the same miRNAs were found in the comparisons of AD versus controls and APOE ε4/4 versus e2/3.

**Fig. 2.**
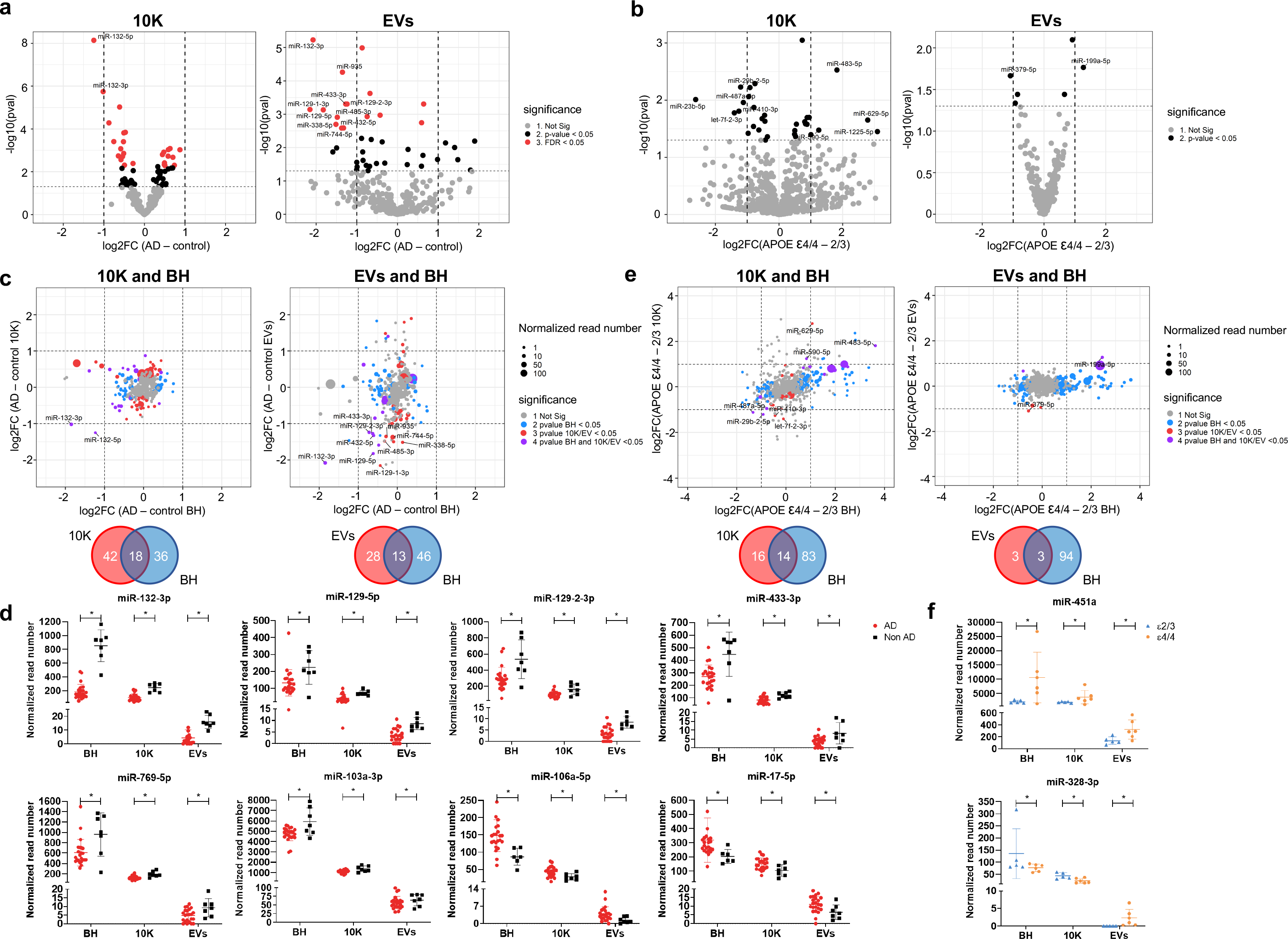
bdEV miRNAs with differential expression in AD. (a) Volcano plots showing 10K (left) and EV (right) miRNA log2 fold changes (Log2FC) and p values (pval) for AD versus control (b) Volcano plots showing 10K (left) and EV (right) miRNA log2 fold changes (Log2FC) and p values (pval) for APOE ε4/4 and APOE ε2/3 carriers. (a) and (b): Thresholds for two-fold change and p value < 0.05 are indicated by dashed lines. Significant changes are indicated with different colors. Grey: non-significant (Not Sig), black: non-adjusted p value < 0.05, and red: FDR < 0.05. (c) miRNA log2 fold changes (Log2FC) for AD and control for BH were plotted against 10K (upper left) and EVs (upper right). Venn diagrams of miRNAs differentially expressed between AD and controls in 10K (bottom left) versus BH, and EVs versus BH (bottom right). (d) The normalized abundance of eight miRNAs dysregulated in AD and control in common in BH, 10K, and EV fractions. Data are presented as mean +/- SD. (e) miRNA log2 fold changes (Log2FC) between APOE ε4/4 and APOE ε2/3 in BH were plotted against 10K (upper left) and EVs (upper right). Venn diagrams of miRNAs differentially expressed between APOE ε4/4 and APOE ε2/3 in 10K (bottom left) versus BH, and EVs versus BH (bottom right). (c) and (e): Dashed lines indicate log2FC of one (up or down). Colored dots indicate miRNAs with p values < 0.05 in BH (blue), in 10K/EV (red), or in both BH and 10K/EVs (purple), or unchanged transcripts (grey). Dot size represents the mean normalized abundance of individual miRNAs. (f) The normalized abundance of two miRNAs dysregulated in APOE ε4/4 vs APOE ε2/3 in BH, 10K, and EV fractions. Data are presented as mean +/- SD.

**Table 2.**
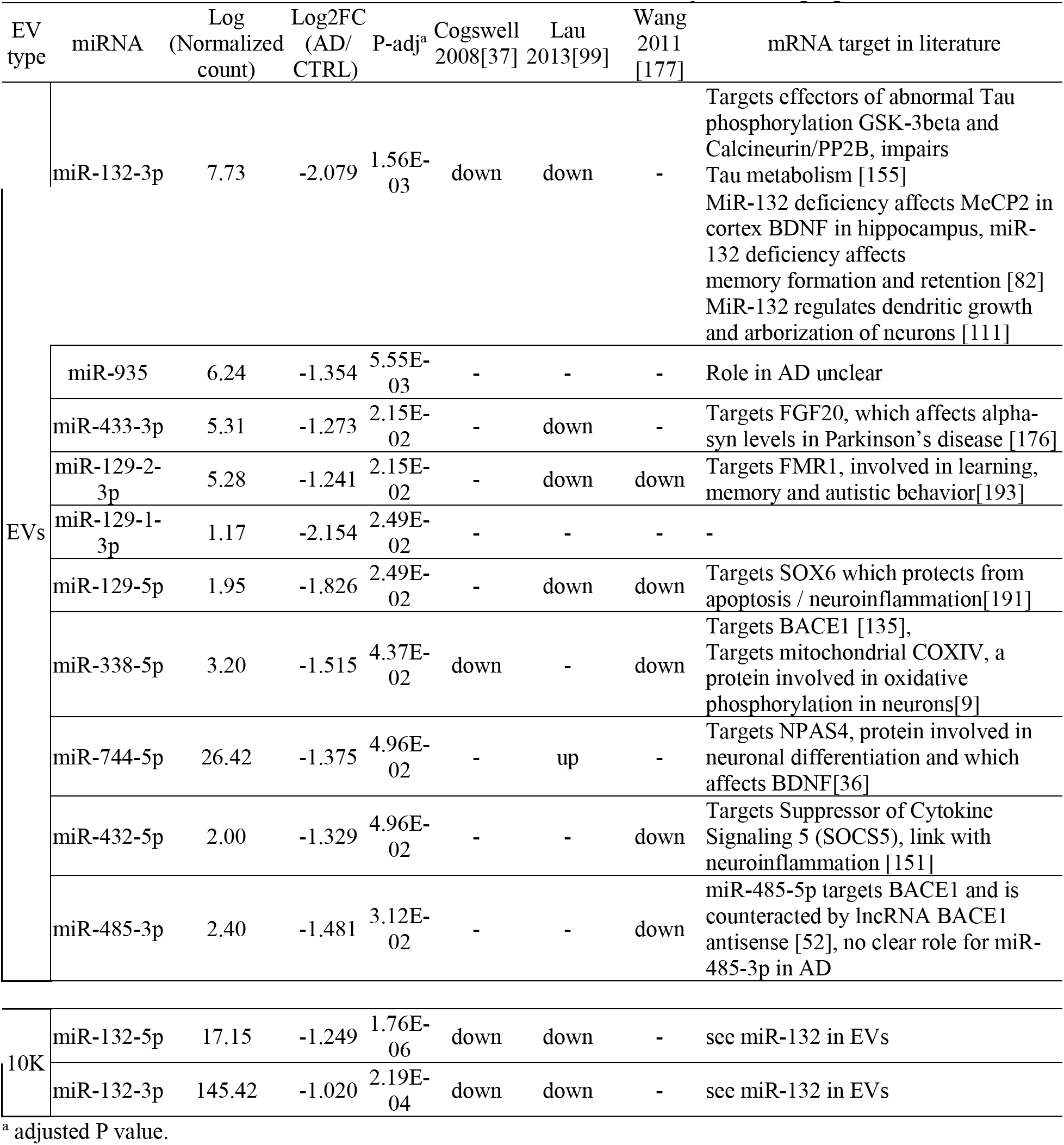
Significantly different miRNAs (adjust P value < 0.05) with twofold change between AD and control in EVs and 10K. Up/downregulation of these EV miRNAs was compared with brain miRNA change in three earlier studies. Studies that have identified involvement of these miRNAs in AD, and their potential target genes are listed.

**Table 3.**
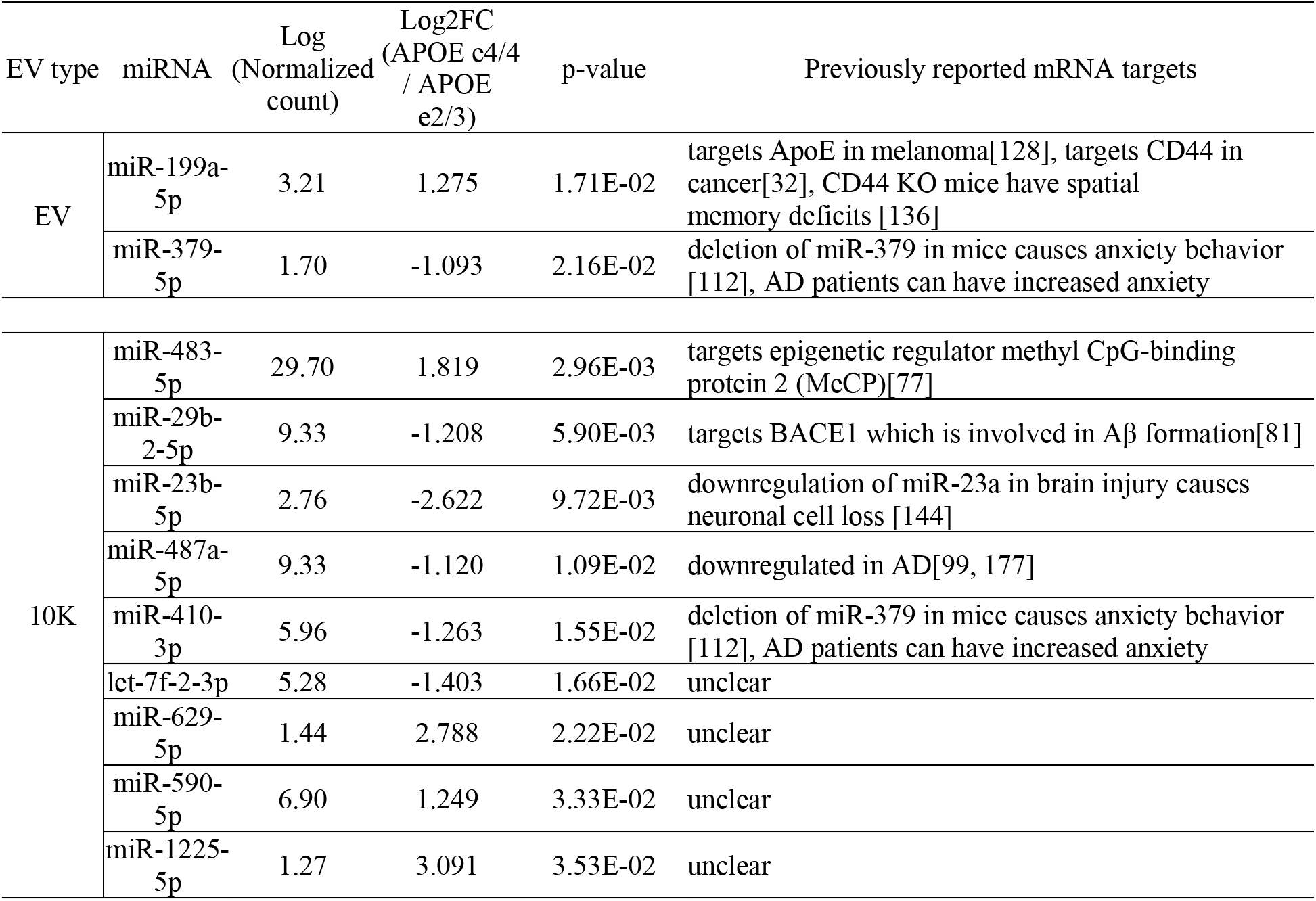
Significantly different miRNAs (adjusted P value < 0.05) with twofold change between APOE e4/4 and e2/3 carrier in EVs and 10K. Up/downregulation of these EV miRNAs was compared with brain miRNA change in three earlier studies. Studies that have identified involvement of these miRNAs in AD, and their potential target genes are listed.

To assess whether AD-associated miRNA differences in the extracellular space reflect overall changes in brain tissue, we compared miRNA fold-changes of 10K and EVs with BH. We highlighted miRNAs that were differentially abundant only in BH (blue dots, Fig. 2c), only in 10K or EV (red dots), or in both EVs and BH (purple dots). Around 30% of differentially abundant EV miRNAs reflected differences that could also be observed in BH (18 miRNAs in 10K and 13 in EV, Fig. 2c). miRNAs that showed robust differential abundance (FDR < 0.05 and log2 fold-change > 2) were found within this subset of miRNAs. Eight miRNAs showed consistent changes across BH, 10K, and EVs (Fig. 2d). Analyzing the higher- and lower-risk APOE genotypes as above, more than 90 miRNAs were differentially abundant in BH (blue dots), while relatively few miRNAs differed in EVs (red dots). The 10K fraction contained more differentially incorporated miRNAs than EVs. Of the differentially abundant miRNAs, 14 showed consistent change between 10K and BH, but only three between EVs and BH (Fig. 2e). The expression levels of only two miRNAs appeared to be consistent across BH, 10K, and EVs (Fig. 2f). Curiously, one of these was the red blood cell-specific miR-451a.

### Differential expression of other non-coding RNA biotypes in brain tissue and EVs related to AD pathology

Since incorporation into EVs of non-miRNA non-coding RNAs can be modulated by external stimuli imposed on cells [33, 46], and since these ncRNAs may also contribute to intercellular communication [72, 122], we examined individual snoRNAs, snRNAs, Y-RNAs, and tRNAs. snoRNAs were relatively depleted from EVs (Fig. S2d), and we did not find any significant differences in snoRNA content of BH, 10K or EVs in AD versus controls (data not shown). Additionally, no differences in snRNA were found for 10K or EVs in AD versus controls, although a significant upregulation of RNU4-2 in AD brain homogenates was observed (Fig. S5a). We also observed slight differences in Y-RNA content of AD versus control EVs (Fig. S5b). Compared with non-AD controls, 10K fractions of AD patients had higher levels of Y4-RNA (p value < 0.05), while EVs contained lower levels of Y1-RNA (p value < 0.05).

Interestingly, we found many differences in tRNA content of EVs in AD versus controls (Fig. 3a). Most of these differences were subtle, however, as none of the tRNAs were enriched more than twofold. In EVs, we observed changes in various isodecoders of tRNA-Cys-GCA and tRNA-Gly-GCC. Additionally, we observed changes in tRNA-Val, tRNA-Arg, tRNA-Pro, and different tRNA-Ala isoacceptors. In contrast, no clear differences in tRNA content were found in 10K. Next, we compared the tRNA content of 10K and EVs from APOE ε4/4 versus ε/3 carriers (Fig. 3b). The fold-differences in tRNA dependent on APOE genotype were larger than those dependent on AD status, although fewer significant tRNAs were found. Two tRNA-Gly-CCC isodecoders were enriched in 10K from APOE ε2/3 carriers, while no significant differences were observed in EVs (as defined by FDR < 0.05).

**Fig. 3.**
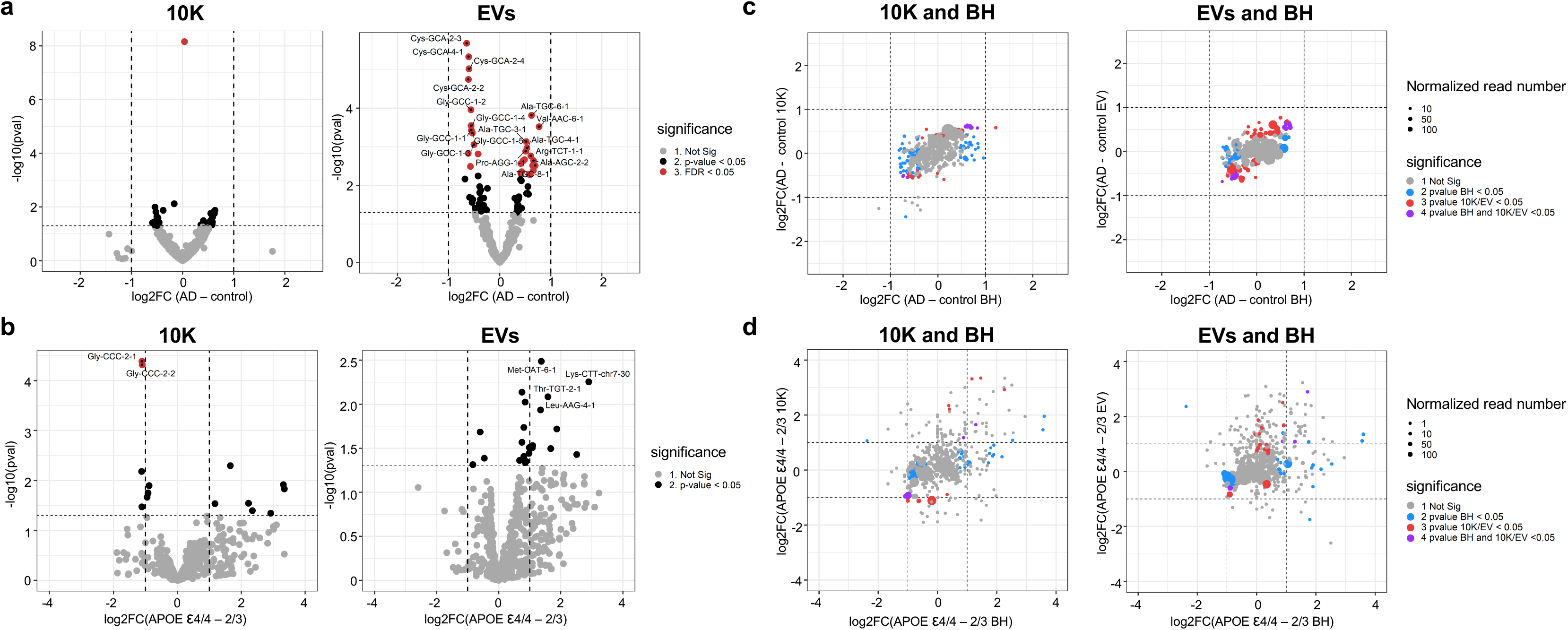
bdEV tRNAs with differential expression in AD. (a) Volcano plots showing 10K (left) and EV (right) tRNA log2 fold changes (Log2FC) and p values (pval) for AD versus control (b) Volcano plots showing 10K (left) and EV (right) tRNA log2 fold changes (Log2FC) and p values (pval) for APOE ε4/4 versus APOE ε2/3. (a) and (b): Thresholds for two-fold change and p value < 0.05 are indicated by dashed lines. Significant changes are indicated with different colors. Grey: non-significant (Not Sig), black: non-adjusted p value < 0.05, and red: FDR < 0.05. (c) tRNA log2 fold changes (Log2FC) for AD and control for BH were plotted against 10K (left) and EVs (right). (d) tRNA log2 fold changes (Log2FC) between APOE ε4/4 and APOE ε2/3 carriers for BH were plotted against 10K (left) and EVs (right). (c) and (d): Dashed lines indicate log2FC of one (up or down). Colored dots indicate tRNAs with p values < 0.05 in BH (blue), in 10K/EV (red), or in both BH and 10K/EVs (purple), or unchanged transcripts (grey). Dot size represents the mean normalized abundance of individual tRNAs.

We subsequently compared the differences in EV-associated tRNA with differences in brain homogenates (Fig. 3c). As for miRNAs, most differences were observed only in the brain homogenates or in EVs, and only a small number of tRNAs differed in both. Comparing APOE genotype-dependent differences in the tRNA content of EVs and BH, we observed large differences only in BH or only in EVs, but only a small number of tRNAs that differed in both (Fig. 3d).

### Proteomics and regulatory pathways

Analysis of data collected by label-free mass spectrometry identified 1675, 801, and 1197 proteins in BH, 10K, and EVs (Supplemental Table 3), respectively. Most proteins were identified in both AD and control in BH (73.6%), 10K (66.5%), and EVs (66.6%) (Fig. 4a, Fig. S6a). Enrichment of common and AD “unique” proteins in BH, 10K, and EVs according to presumed regulatory pathways as identified in the Kyoto Encyclopedia of Genes and Genomes (KEGG) database was observed for pathways directly or indirectly related to neurodegenerative diseases (Fig. 4b, Fig. S6b). Metabolic pathways (e.g., carbon metabolism, glycolysis, and oxidative phosphorylation) were widely enriched for common and AD-unique proteins. Proteins with known involvement in Alzheimer’s disease, Parkinson’s disease, and Huntington’s disease were enriched in AD-unique proteins in 10K and EVs (Fig. 4b) and common proteins in BH (Fig. S6b). Principal component analysis (PCA) of the EV proteome content showed a separation of AD and control groups (Fig. 4c, right). In contrast, no such separation was observed for 10K (Fig. 4c, left) or BH (Fig. S6c).

**Fig. 4.**
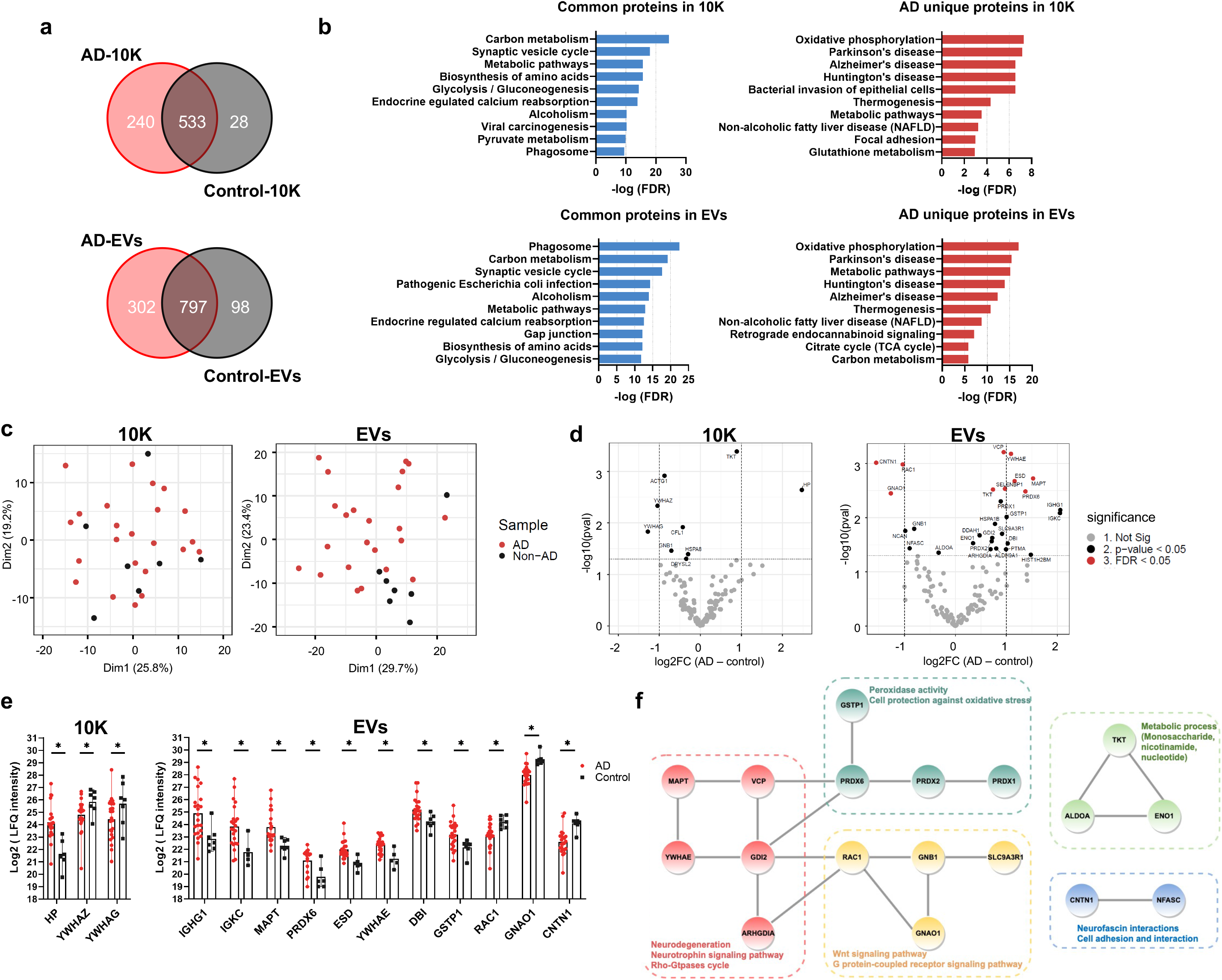
bdEV protein content reflects differences between AD and control brain tissues. (a) Venn diagrams of proteins identified in 10K and EVs from AD and control patients. (b) Top 10 pathways ranked by FDR-corrected p value of 10K and EV proteins according to the Kyoto Encyclopedia of Genes and Genomes (KEGG). (c) Principal component analysis (PCA) based on proteome content of 10K (left) and EVs (right). (d) Volcano plots showing 10K (left) and EV (right) protein log2 fold changes (Log2FC) and p values (pval) for AD versus control. Thresholds for two-fold change and p value < 0.05 are indicated by dashed lines. Significant changes are indicated with different colors. Grey: non-significant (Not Sig), black: non-adjusted p value < 0.05, and red: FDR < 0.05. (e) Expression level of proteins differentially expressed between AD and control with fold change > 2 (non-adjusted p value < 0.05) in 10K (left) and EVs (right). Data are presented as mean +/- SD. (f) STRING protein interaction network analysis indicated that 18 proteins (out of 31 proteins showing differential expression (non-adjusted p value < 0.05) between AD and control in EVs) were enriched with known high confidence (0.7 on a scale of 0-1) protein-protein interactions. Protein clusters are indicated with different colors based on predicted functions.

### Differentially abundant proteins: AD versus control

Label-free quantitation (LFQ) was used to identify up- and downregulated proteins in AD (Fig. 4d and Fig. S6d). More proteins were up- or downregulated in AD in the EV fraction (Fig. 4d, right) than in 10K (Fig. 4d left) or BH (Fig. S6d). EVs have increased potential to indicate the difference between AD and controls as compared with 10K and BH. Three proteins (10K) and 11 proteins (EVs) differed by more than two-fold between AD and control (Fig. 4e, S7a-b). Examining proteins that were differentially abundant in EVs by Protein-Protein Interaction Networks Functional Enrichment Analysis (STRING), 18 out of 31 had high protein to protein interaction confidence scores (0.7 on a scale of 0-1), participating in AD-related processes such as neurodegeneration, neurotrophin signaling, oxidative regulation, and metabolic regulation (Fig. 4f). Prominent among these was microtubule-associated protein tau (MAPT), which forms neurofibrillary tangles in AD brains.

We next measured the concentration of total Tau and Tau with phosphorylated threonine 231 (phosTau) in lysed BH, 10K, and EVs by ECL-immunoassay in two independent clinical cohorts. Normalized to total protein input, total Tau and phosTau were significantly increased in bdEVs of AD versus controls in both cohort 1 (Fig. 5a) and cohort 2 (Fig. 5b), while in the 10K fraction only phosTau increased in AD and only in cohort 2 (Fig. 5b). In BH, phosTau was significantly increased in AD compared with controls while Tau did not change in either cohort (Fig. 5a-b). phosTau increased more than 12-fold in AD EVs compared with controls, while Tau was increased around 2-fold (Fig. 5c). To test the predictive power of Tau and phosTau in BH, 10K and EVs, receiver operating characteristic (ROC) curves were generated by logistic regression (Fig. 4d). Total Tau level in EVs was significant to distinguish AD from control with an area-under-curve (AUC) of 0.67±0.08, while phosTau levels in BH and EVs more accurately distinguished AD from control, reflected by an AUC of 0.93±0.03 and 0.90±0.014, respectively. In contrast, neither Tau or phosTau in 10K distinguished AD from control significantly. To assess whether changes in Tau or phosTau in EVs or 10K reflected those in BH, we determined their correlations. No strong correlation was observed for total tau and phosTau levels (BH vs 10K or EVs; Fig. S8) in AD patients, while phosTau levels in BH and EVs were positively correlated in controls (R=0.73, p=0.00088; Fig. S8). Further verification of proteomics data was also conducted on PRDX1, PRDX6, ENO2, and HSP70 by ECL-immunoassay, however, no significant changes were found consistent between two cohorts after protein input normalization (data not shown).

**Fig. 5.**
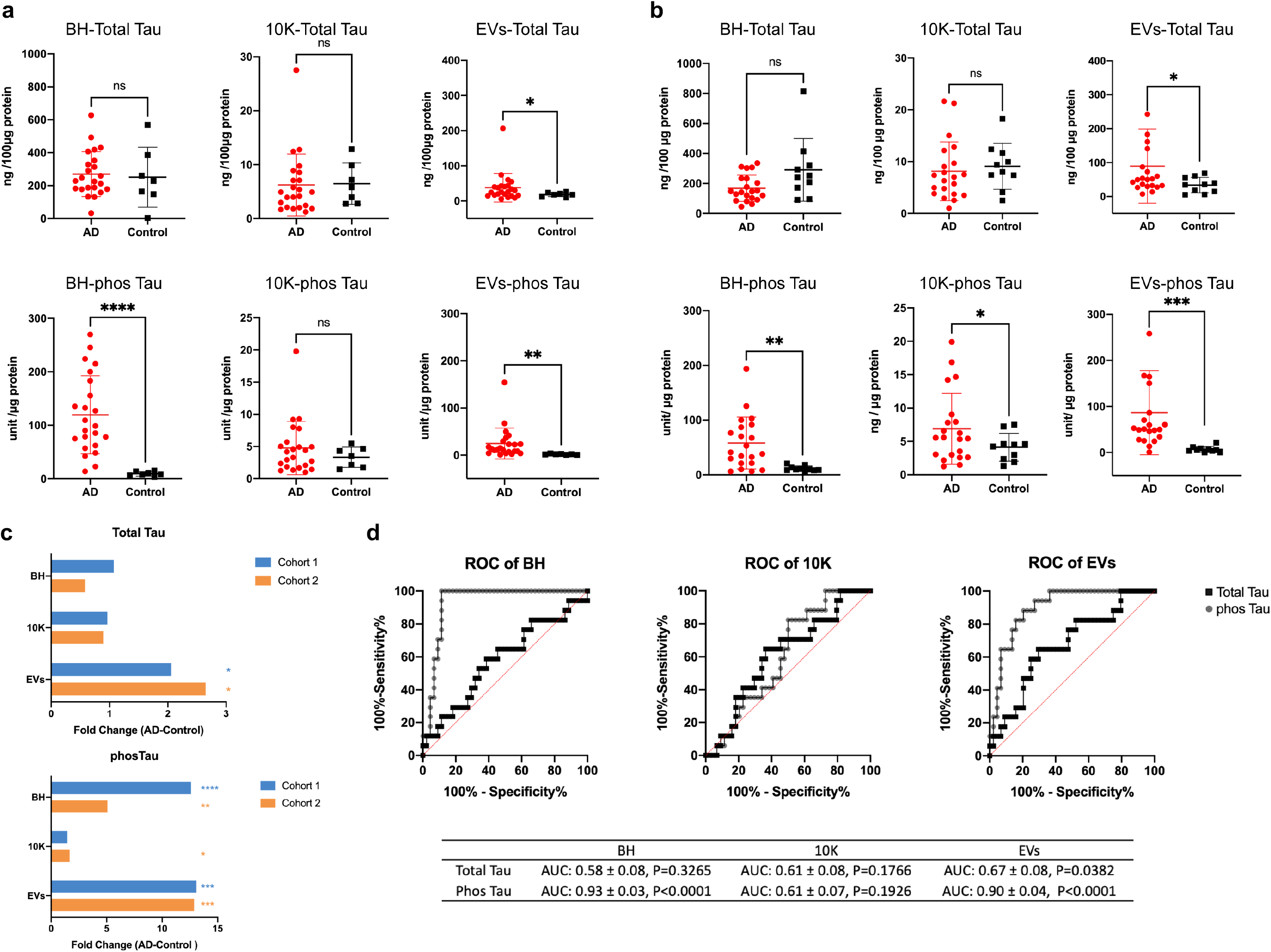
Higher total tau and phosphorylated tau protein levels in bdEVs from AD patients compared with controls. (a) (b) Levels of total tau (per 100 µg protein input) and phosphorylated tau (phosTau) (per 1 µg protein input) protein in BH, 10K, and EVs from cohort 1 (A) and cohort 2 (B), measured by ECL immunoassay. Data are presented as mean +/- SD. ns: no significant difference (p > 0.05), *p ≤ 0.05, **p ≤ 0.01, ***p ≤ 0.001, ****p ≤ 0.0001 by two-tailed Welch’s t-test. (c) Tau and phosTau fold-change in AD compared to control patients. (d) Receiver operating characteristic (ROC) curves are presented for Tau and phosTau in BH, 10K, and EVs from cohort 1 and 2 patients. The area under curve (AUC) with 95% CI and p value are shown.

### Proteomics: APOE ε4/4 and APOE ε2/3

Next, we assessed differences in the proteome between APOE ε4/4 and ε2/3 genotypes (AD samples only). Most proteins were detected in more than 50% of individuals in each group (i.e., n > 3 in each; Fig. 6a, Fig. S9a). Most proteins were found in common in the ε4/4 and ε2/3 groups (71.4% in 10K, 70.3% in EVs, and 81.5% in BH). KEGG pathway analyses of APOE ε4/4-“unique” proteins in 10K and EVs corresponded with metabolism-related pathways (carbon, glutathione, and galactose metabolism) (Fig. 6b). KEGG pathway analysis of APOE ε2/3-“unique” proteins in 10K and EVs did not return enriched pathways (data not shown), although numerous pathways were enriched in BH of APOE ε2/3 carriers (Fig. S9b). Moreover, both ε4/4 and ε2/3 contained enriched proteins in BH that were involved in Alzheimer’s disease, Huntington’s disease, and oxidative phosphorylation (Fig. S9b).

**Fig. 6.**
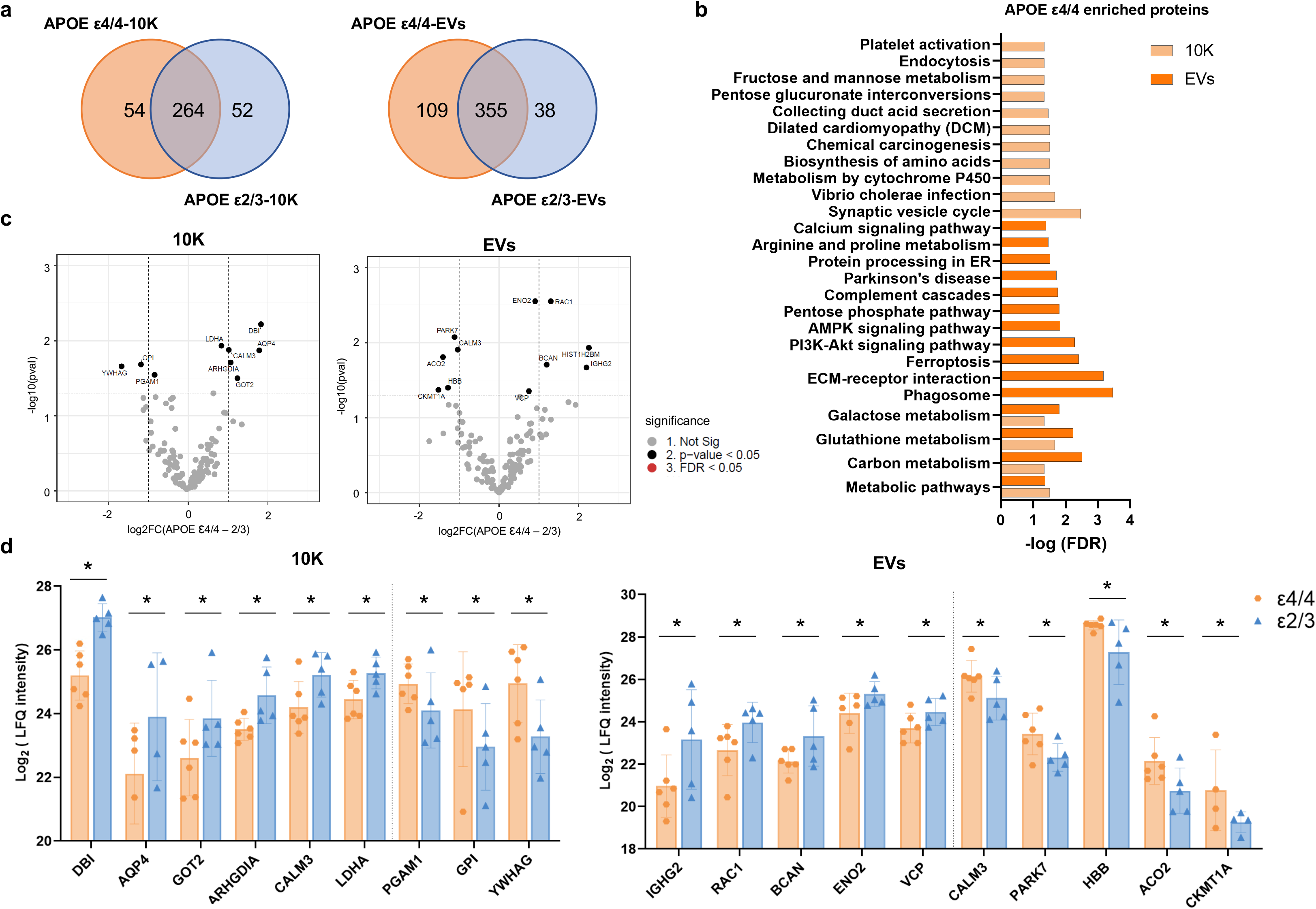
bdEV proteins with differential expression in APOE ε4/4 and APOE ε2/3 carriers. (a) Venn diagrams of proteins identified in 10K and EVs from APOE ε4/4 and APOE ε2/3 carriers. (b) Significant pathways of APOE ε4/4 10K and EV proteins according to the Kyoto Encyclopedia of Genes and Genomes (KEGG). (c) Volcano plots showing 10K (left) and EV (right) protein log2 fold changes (Log2FC) and non-adjusted p values (pval) between APOE ε4/4 and APOE ε2/3 carriers. Thresholds for two-fold change and p value < 0.05 are indicated by dashed lines. Significant changes are indicated with different colors. Grey: non-significant (Not Sig), black: non-adjusted p value < 0.05. (d) Expression level of proteins differentially expressed between APOE ε4/4 and APOE ε2/3 in 10K (left) and EVs (right). Data are presented as mean +/- SD.

Via differential expression analysis, we identified several proteins that were upregulated or downregulated in ε4/4 vs ε2/3 genotypes (Fig. 6c (10K and EVs) and Fig. S9c (BH)). The expression levels of dysregulated proteins between ε4/4 vs ε2/3 are shown in Fig. 6d and Fig. S10a-b. We observed more dysregulated proteins in the 10K and EV fractions than in BH. The potential functions of these proteins in neurodegenerative disease progression are summarized in Table 4 and 5.

**Table 4.** The potential function of significantly different proteins (P value < 0.05) between APOE e4/4 and e2/3 carrier in EVs.

| Protein symbol | Protein name | Enriched in | Functions | References |
| --- | --- | --- | --- | --- |
| RAC1 | Ras-related C3 botulinum toxin substrate 1 | $\epsilon$ 2/3 | Rho-GTPases, membrane remodeling | [2, 19, 166] |
| BCAN | Brevican core protein | $\epsilon$ 2/3 | neurite outgrowth, stabilizes synapses | [12, 84] |
| ENO2 | γ-enolase | $\epsilon$ 2/3 | neurotrophic and neuroprotective | [73, 74, 78] |
| VCP | Transitional endoplasmic reticulum ATPase | $\epsilon$ 2/3 | mitochondrial mitophagy impairment, abnormal protein aggregate and clearance | [71, 184] |
| CALM3 | Calmodulin-3 | $\epsilon$ 4/4 | cytosolic $\text{Ca}^{2+}$ concentration regulation | [86] |
| PARK7 | Protein deglycase DJ-1 | $\epsilon$ 4/4 | antioxidant defense | [13, 34] |
| ACO2 | Aconitate hydratase | $\epsilon$ 4/4 | energy metabolism, mitochondrial function | [13, 87, 107, 131] |
| CKMT1A | Creatine kinase U-type | $\epsilon$ 4/4 | ATP production | [13, 156] |
| HBB | Hemoglobin subunit beta | $\epsilon$ 4/4 | oxygen homeostasis regulation | [149, 152] |

**Table 5.** The potential function of significantly different proteins (P value < 0.05) between APOE e4/4 and e2/3 carrier in 10K.

| Protein symbol | Protein name | Enriched in | Function in CNS disease | References |
| --- | --- | --- | --- | --- |
| DBI | Acyl-CoA-Binding Protein | $\epsilon 2/\epsilon 3$ | GABA receptor modulation; neurogenesis | [4, 48, 68] |
| GOT2 | Glutamate oxaloacetate transaminase, mitochondrial | $\epsilon 2/\epsilon 3$ | mitochondrial energy metabolism | [8] |
| ARHGDIA | Rho GDP-dissociation inhibitor 1 | $\epsilon 2/\epsilon 3$ | modulation of GTPases; cell motility regulation | [126] |
| CALM3 | Calmodulin-3 | $\epsilon 2/\epsilon 3$ | cytosolic $\text{Ca}^{2+}$ concentration regulation | [86] |
| LDHA | L-lactate dehydrogenase A chain | $\epsilon 2/\epsilon 3$ | glycolysis | [94] |
| PGAM1 | Phosphoglycerate mutase 1 | $\epsilon 4/\epsilon 4$ | glycolysis metabolism; cell proliferation; | [90, 183] |
| GPI | Glucose-6-phosphate isomerase | $\epsilon 4/\epsilon 4$ | glycolysis metabolism; neuroleukin function | [53, 56, 116] |
| YWHAG | 14-3-3 protein gamma | $\epsilon 4/\epsilon 4$ | neuronal differentiation | [38, 95] |

## DISCUSSION

The roles of EVs in regulating CNS diseases have been inferred predominantly from studies of *in vitro* models and biofluid EVs, with growing but still limited study of tissue EVs. Here, we compared protein and small RNA contents of brain homogenate with those of a “10K” pelleted extracellular fraction and a purified EV fraction of control and late-stage AD brain, including several APOE genotypes. Although the concentrations of recovered particles did not differ significantly with AD pathology, several bdEV miRNAs, tRNAs, and proteins were dysregulated not only between AD and controls, but also between the APOE genotypes representing the most “distant” risk groups in our study, the ε4/4 and ε2/3 carriers. Proteome differences between AD and controls were most pronounced for EVs, suggesting that EV proteins may have the best biomarker potential, at least for late-stage AD. The dysregulated molecules identified in our study, especially those involved in aging and neurodegeneration pathways, may be involved in CNS disease mechanisms and constitute new biomarkers for disease monitoring.

### Does AD or APOE genotype affect EV production or subtypes?

bdEVs are released as a mixture of various subtypes and from diverse cells, and both biogenesis and cargo loading of specific EV subtypes may be affected by AD. Indeed, we observed a shift towards recovery of smaller particles from AD brain, and our finding of more APOE genotype-related changes in EV fractions suggests that APOE may be more likely to affect cargo loading rather than overall protein expression levels in tissue. Even so, like several previous studies of human cortex bdEVs [59, 120], we did not observe differences in overall EV recovery. Although the APOE ε4 allele is reported to be involved in neuronal, endosomal, and lysosomal system dysfunction [27], we also did not see particle count differences associated with APOE genotype. This contrasts with an earlier report of reduced EV counts in the presence of the APOE ε4 allele [129]. This may be due to our examination of only late-stage AD samples (Braak stage 5-6, CERAD B-C). Previously, MHC class I bdEVs were upregulated in preclinical AD patients compared with controls and late-stage AD [59], and ε4-associated EV differences were found in non-diseased brain tissues of human and mouse [129]. Thus, effects of APOE alleles may measurably affect EVs and their influence on brain function only in young brain and before the onset of late-stage disease [26, 105]. Further studies of APOE and EVs will be needed in young brain and during early disease stages.

### Altered bdEV miRNA expression in AD

RNA sequencing revealed several differentially expressed (DE) miRNAs in AD bdEVs with potential involvement in AD. Importantly, these DE miRNAs largely overlapped with results of earlier studies on miRNAs in AD brain tissues [37, 99, 177]. For example, miRNA-132-3p was downregulated in AD EVs. It was previously found to be downregulated in late-onset AD, negatively correlated with Braak stages and formation of tau tangles in the prefrontal cortex [99]. miR-132 promotes neurite outgrowth and synapse formation [110, 111], with expression driven by the neurotrophin-responsive transcription factor CREB [174]. Additionally, miR-132 knockout mice express increased levels of phosphorylated tau [155] and display memory deficits [82]. In line with downregulated miR-132-5p, we found that tau levels were increased in AD EVs. Downregulated miR-338-5p targets beta-secretase 1 (BACE1) [135], which together with γ-secretase cleaves APP into aggregate-forming Aβ peptides [76]. Downregulated miR-129-5p and miR-432-5p regulate neuroinflammation [151, 191], and miR-129-3p targets fragile X mental retardation 1 (FMR1), which controls migration of cortical neurons [182, 193]. Although our study predominantly highlights downregulated miRNAs, upregulated miRNAs have also been reported in AD bdEVs [31]. We found at least three upregulated miRNAs in common with that study: miR-483-5p, miR-152-3p, and miR-30a-5p [31]. Beyond these validations, differences between the studies may be explained by use of different brain regions for bdEV isolation, as well as differences in separation methods, possibly affecting purity and recovery of EV subtypes. Overall, we found various dysregulated miRNAs in AD EVs that appear to be involved in AD hallmarks such as APP and tau processing, neuronal function, memory formation, and neuroinflammation.

### bdEV miRNAs and APOE genotype

In contrast with AD versus control comparisons, EV miRNA profile differences by APOE were not as pronounced. Nevertheless, several EV-associated miRNAs differed. In the 10K fraction of APOE ε4/4 carriers, miR-29b-5p was downregulated. This miRNA targets BACE, which contributes to Aβ aggregation (Hebert et al PNAS 2008). Additionally, we observed downregulation of miR-379-5p and miR-410-3p, which have been previously implicated in anxiety behavior [112], miR-23a-5p, which is linked to neuronal cell loss [144], and miR-483-5p, which is involved in neuronal development [77]. Our findings are limited, however, since only late-stage carriers were examined, with late Braak stage and similar CNS pathology. miRNAs are quite possibly involved in APOE genotype interactions much earlier in AD, perhaps even in young, putatively healthy individuals. More research is thus needed to address the involvement of these miRNAs in AD pathogenesis.

### AD-dependent differences in additional bdEV-associated ncRNAs

AD-dependent differences in bdEV-associated ncRNAs such as tRNAs and Y-RNAs were also apparent. These ncRNAs are highly abundant in EVs from various sources [15, 46, 65, 125, 165, 175], but their roles in health and disease remain largely unexplored. We found various AD-dependent differences in tRNA content of EVs, as well as APOE-dependent differences in tRNAs in the 10K fraction. Changes in cellular tRNA abundance have been implicated in neurological disorders [96, 164], and dysregulation of cellular tRNA levels affects protein expression in cancer [67]. Interestingly, tRNA fragments released with EVs may modulate gene expression in recipient cells [60]. Small changes in bdEV-associated Y-RNAs (Y1 and Y4) were also observed. Y-RNA is abundantly detected in EVs from various biological sources [47]. Cellular Y-RNA serves as a scaffold for RNA-binding proteins [17]. In AD patients, dysregulation of Y3-RNA binding to enhancer protein HuD (ELAVL4) was shown to cause alternative splicing in neurons [148]. Additionally, EV-associated Y4-RNA activated endosomal RNA sensor TLR7 in monocytes [72]. In brain tissue of AD patients, we observed a significant difference in U4 RNA, which has not been previously reported in AD. As part of the spliceosome complex, U4 is involved in pre-mRNA splicing [113]. In the prefrontal cortex of AD brains, snRNA U1 was found to be upregulated and to accumulate in tau plaques [10, 75]. Upregulation of U1 was additionally implicated in aberrant mRNA splicing and altered APP expression [10]. Our findings encourage further investigations into the role of extracellular non-miRNA ncRNAs in AD and other neurological conditions.

### Implications of bdEV-associated proteins in AD

#### bdEV-associated tau proteins

EVs have been widely reported to propagate misfolded protein MAPT (tau) in neurodegenerative disease [11, 22, 40, 134]. Consistent with other bdEV studies [11, 120, 134], we found a higher level of tau and phosphorylated tau at threonine 231 in EVs of AD brain compared with controls, adding more evidence to EV-mediated tau propagation. However, the mechanism of tau loading into EVs remains unclear. Further study is needed to decide if EVs act as a protective factor to clear tau from the CNS to the periphery, or as a neurotoxic factor to spread tau between neurons and glia, or perhaps some as-yet incompletely understood balance between the two extremes.

#### bdEV-associated antioxidant proteins

Upregulated members of the antioxidant peroxiredoxin (PRDX) family in AD EVs include PRDX1, PRDX2, and PRDX6. Oxidative damage is often implicated in the progression of neurodegenerative diseases. We and our collaborators previously found that PRDXs were highly abundant in EVs derived from both human induced pluripotent stem cells (iPSCs) and mesenchymal stem cells (MSCs), and that these proteins help to reduce oxidative stress in induced senescent cells [106]. Although the protective role of PRDX family members in AD [98, 100, 195] and the association of PRDX with EVs [106, 118, 160] have been reported before, this is the first study to show PRDXs detected in tissue EVs from AD brain. However, our verification of PRDX1 and PRDX6 by ELC immunoassay showed they were only significantly increased in AD versus control in cohort 1 but not in cohort 2 after normalization by protein input (data not shown), which may be caused by the individual variance of patient samples. We also found another antioxidant defense protein, deglucase DJ1 (PARK7), enriched in APOE ε4/4 compared with ε2/3 samples. PARK protects cells against hydrogen peroxide-induced death [91, 150] upon oxidative stress. We hypothesize that this factor may spread via EVs as a neuroprotective response in patients with the risk factor ε4.

#### bdEV-associated mitochondrial and metabolism pathway proteins

Mounting evidence suggests that mitochondrial and metabolism dysfunction contribute to AD by compromising the energy supply [25, 79, 101, 119]. We found numerous bdEV-associated proteins that contribute to mitochondrial and metabolic pathways. The ATPase enzyme member valosin-containing protein/p97 (VCP) was upregulated in AD bdEVs. VCP is thought to bring about mitophagy impairment, leading to mitochondrial dysfunction in AD [71, 184]. Higher levels of tricarboxylic acid cycle (TCA) protein mitochondrial aconitate hydratase (ACO2) were previously found in multiple brain regions of AD patients [190] and here appeared to be enriched in bdEVs of APOE ε4/4 patients compared with ε2/3. We also identified changes in bdEV proteins that are involved in glycolytic and carbon metabolism, including transketolase (TKT) [28, 64], alpha-enolase (ENO1) [23, 132], and fructose-bisphosphate aldolase A (ALDOA) [58]. Further exploration of protein modification and enzymatic activity of these proteins is now indicated to unveil mechanisms of energy metabolism deficits in AD.

#### bdEV-associated proteins in neurogenesis

Impairment of pathways involved in neurogenesis/differentiation are thought to be early clinical events in aging and AD development [18, 35, 41, 101, 102]. Among neurogenesis-related proteins, the 14-3-3 protein family of ubiquitous phosphoserine/threonine–binding proteins is highly abundant in brain and implicated in nervous system development and maintenance [14, 16, 20]. Our study shows dysregulation of 14-3-3 isoforms ε (YWHAE), ζ (YWHAZ), and γ (YWHAG) in the 10K and EV fractions. 14-3-3 proteins indirectly regulate activation of Rho family GTPases [167]. Interestingly, several proteins in the Rho-GTPase cycle were also dysregulated in our bdEVs, including Ras-related C3 botulinum toxin substrate 1/3 (RAC1/3), and Rho GDP-dissociation inhibitors 1 and 2 (ARHGDIA, GDI2). We also found that contactin-1 (CNTN1) and neurofascin (NFASC), two proteins involved in axonal guidance and neuron projection development [49, 63, 70, 153], were downregulated in AD bdEVs, which may reflect neurodegeneration. In addition, some neuroprotective proteins were also found to be downregulated in bdEVs from APOE ε4/4 patients compared to ε2/3, such as γ-enolase (ENO2) [73, 74, 78] and brevican core protein (BCAN) [12, 57, 84].

In summary, this is the first study thoroughly examining bdEV small RNA and protein content from clinical AD patients with different APOE genotypes. Our results identified the composition of RNAs (including miRNAs, tRNAs, and Y-RNAs) and proteins in bdEVs that varied with AD pathology and APOE genotypes. These bdEV-transported molecules may play critical roles in modulating AD progression and thus have potential for exploitation as biomarkers and therapeutic targets.

### Contribution statements

All authors contributed to the study conception and design. Material preparation, data collection and analysis were performed by Yiyao Huang, Tom A. P. Driedonks, Lesley Cheng, Andrey Turchinovich, and Harinda Rajapaksha. Drafts of the manuscript were written by Yiyao Huang, Tom A. P. Driedonks, and Kenneth W. Witwer. All authors commented on previous versions of the manuscript. All authors read and approved the final manuscript.

## Data Availability

Nucleic acid sequencing data have been deposited with the Gene Expression Omnibus, accession GSE159541. We have submitted all relevant details of our experiments to the EV-TRACK knowledgebase (EV-TRACK ID: EV200126). Any other data are available on reasonable request.

https://www.ncbi.nlm.nih.gov/geo/query/acc.cgi?acc=GSE159541

## ACKNOWLEDGMENTS

This work was supported in part by grants from the US National Institutes of Health: AI144997 (to KWW, with support for TAPD), MH118164 and AG057430 (to VM and KWW), by UG3CA241694 (to KWW), supported by the NIH Common Fund, through the Office of Strategic Coordination/Office of the NIH Director, and by the National Health and Medical Research Council of Australia (GNT1132604 to AFH). VM and KWW gratefully acknowledge support from the Richman Family Precision Medicine Center of Excellence in Alzheimer’s Disease including helpful comments and advice from founder and director Constantine Lyketsos. Thanks to: Kenneth Pienta, Johns Hopkins University School of Medicine, for access to the nanoFCM flow nanoAnalyzer platform; Mitchell Science Writing for manuscript editing and citation formatting; and the La Trobe University Comprehensive Proteomics Platform. We also thank members of the Witwer and Retrovirus Laboratories, Johns Hopkins University School of Medicine, and various members of the International Society for Extracellular Vesicles for valuable discussions and support.

---

**Fig. S1.**
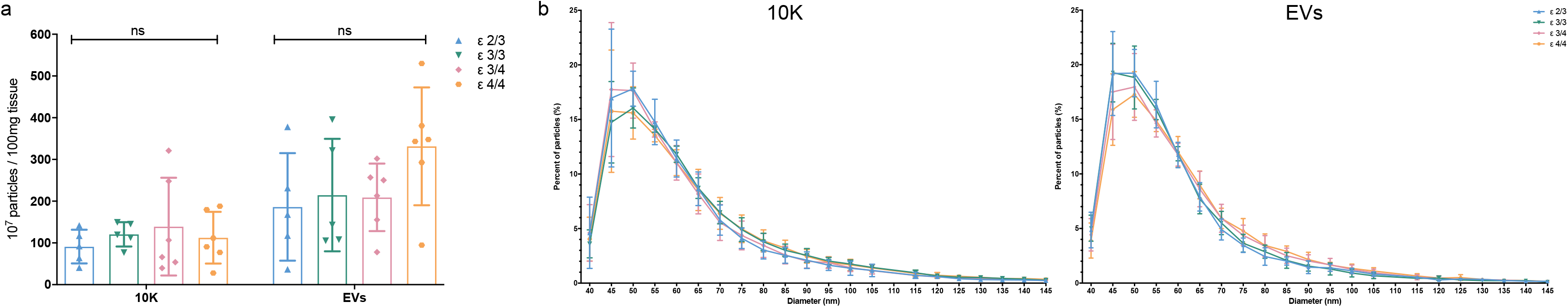
Particle yield and size distribution of bdEVs from AD samples with different APOE genotypes. Particle concentration (a) and size distribution (b) of 10K and EV fractions from AD patients classified by APOE genotype as measured by NFCM. Data are presented as mean +/- SD. ns: no significant difference (p > 0.05) between the two APOE genotype groups by two-tailed Welch’s t-test.

**Fig. S2.**
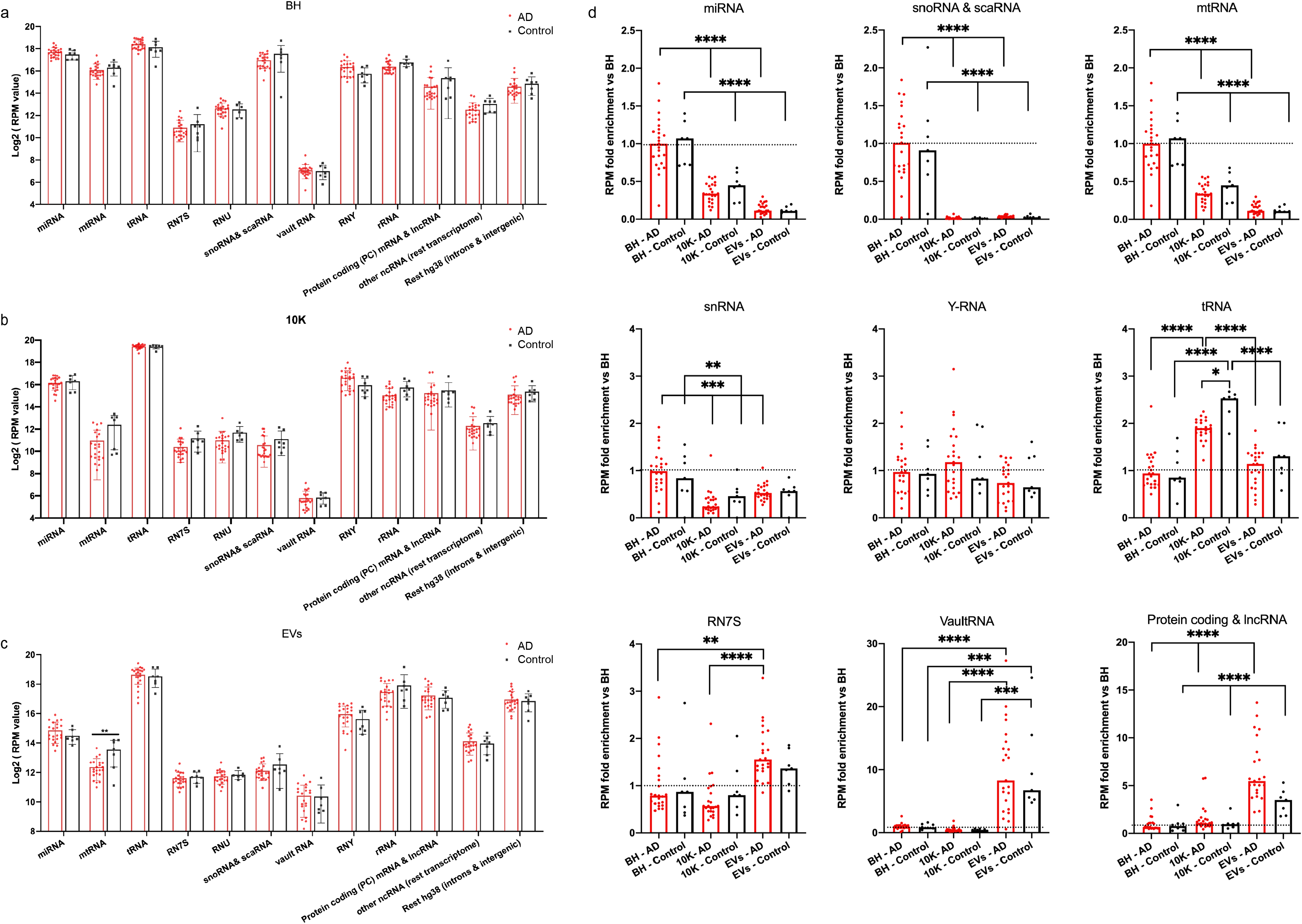
Small RNA biotypes in BH, 10K, and EV fractions from AD and control patients. The log2 reads per million mapped reads (Log2RPM) of small RNA biotypes in BH (a), 10K (b), and EVs (c) of AD and control. Data are presented as mean +/- SD. **p ≤ 0.01 by two-tailed Welch’s t-test. (b) Normalized RNA class read counts in 10K and EVs were scaled to the normalized RNA class read counts in BH. Data are presented as mean +/-SD. *p ≤ 0.05, **p ≤ 0.01, ***p ≤ 0.001, ****p ≤ 0.0001 by two-tailed Welch’s t-test.

**Fig. S3.**
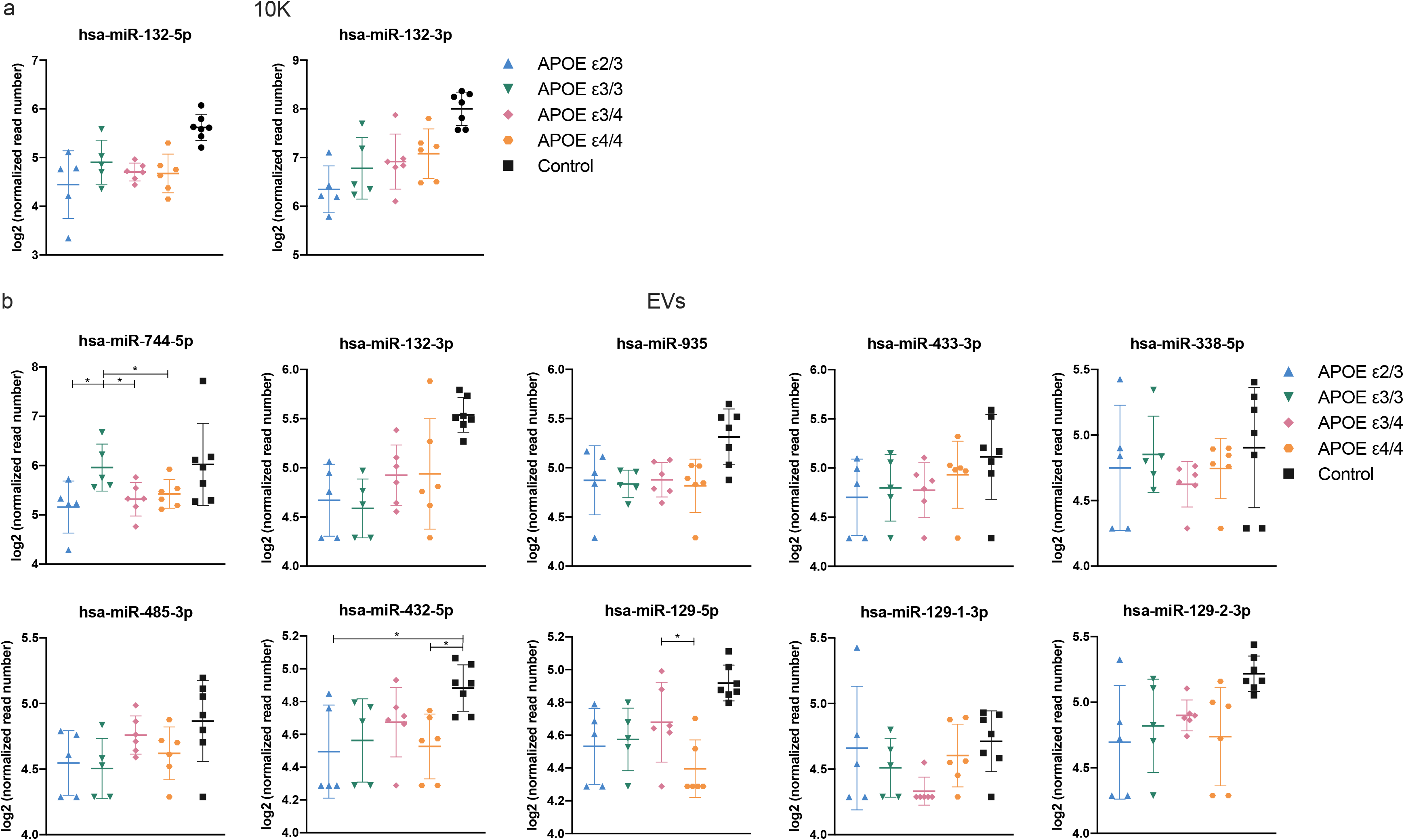
Normalized count of miRNAs differentially expressed between AD and control with fold change > 2 in 10K (a) and EVs (b) of patients classified by APOE genotype. *p value < 0.05 by DeSeq2 differential gene expression analysis based on the negative binomial distribution.

**Fig. S4.**
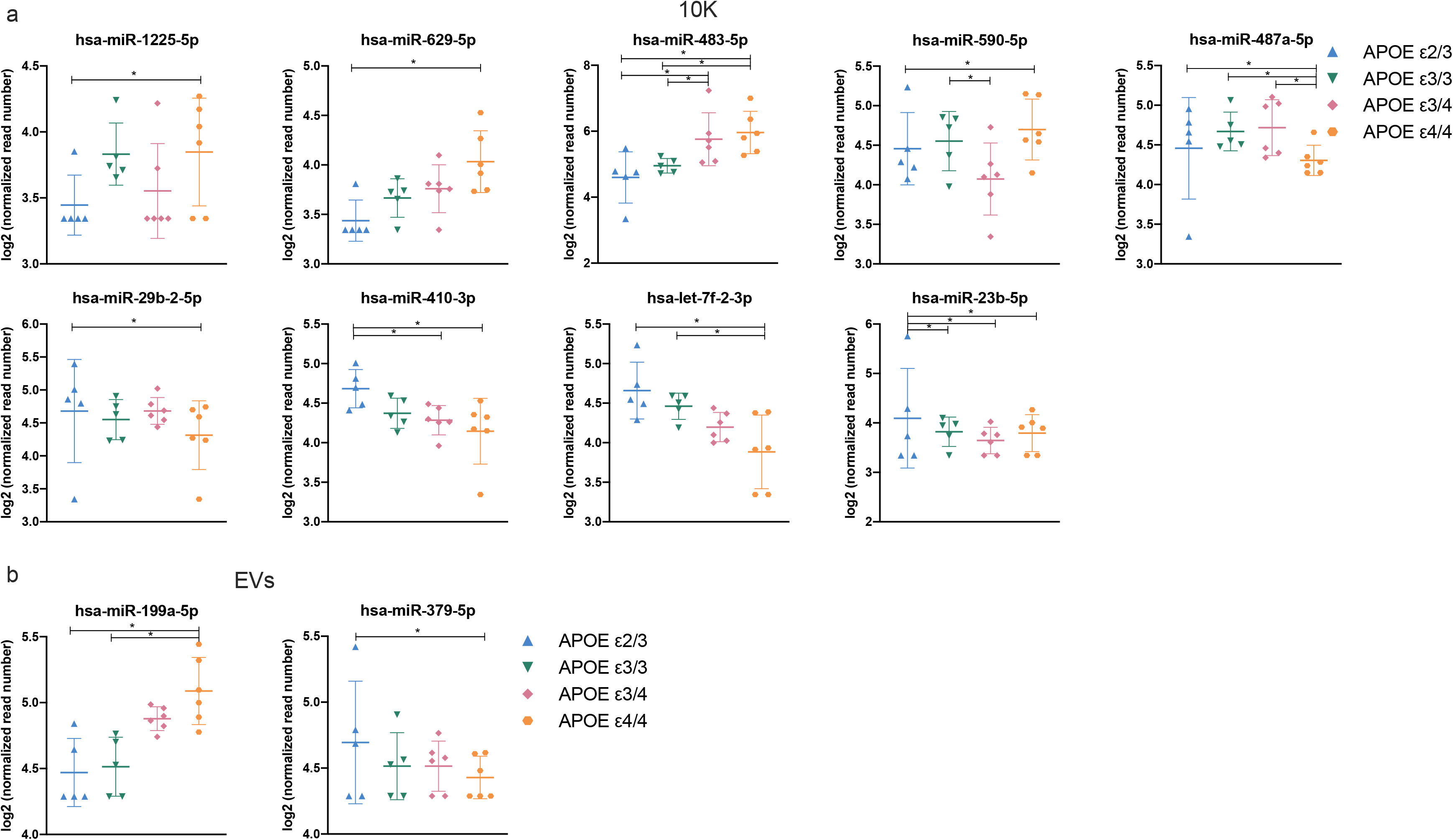
Normalized count of miRNAs differentially expressed in APOE ε4/4 versus APOE ε2/3 carriers with fold change > 2 in 10K (a) and EVs (b). *p value < 0.05 by DeSeq2 differential gene expression analysis based on the negative binomial distribution.

**Fig. S5.**
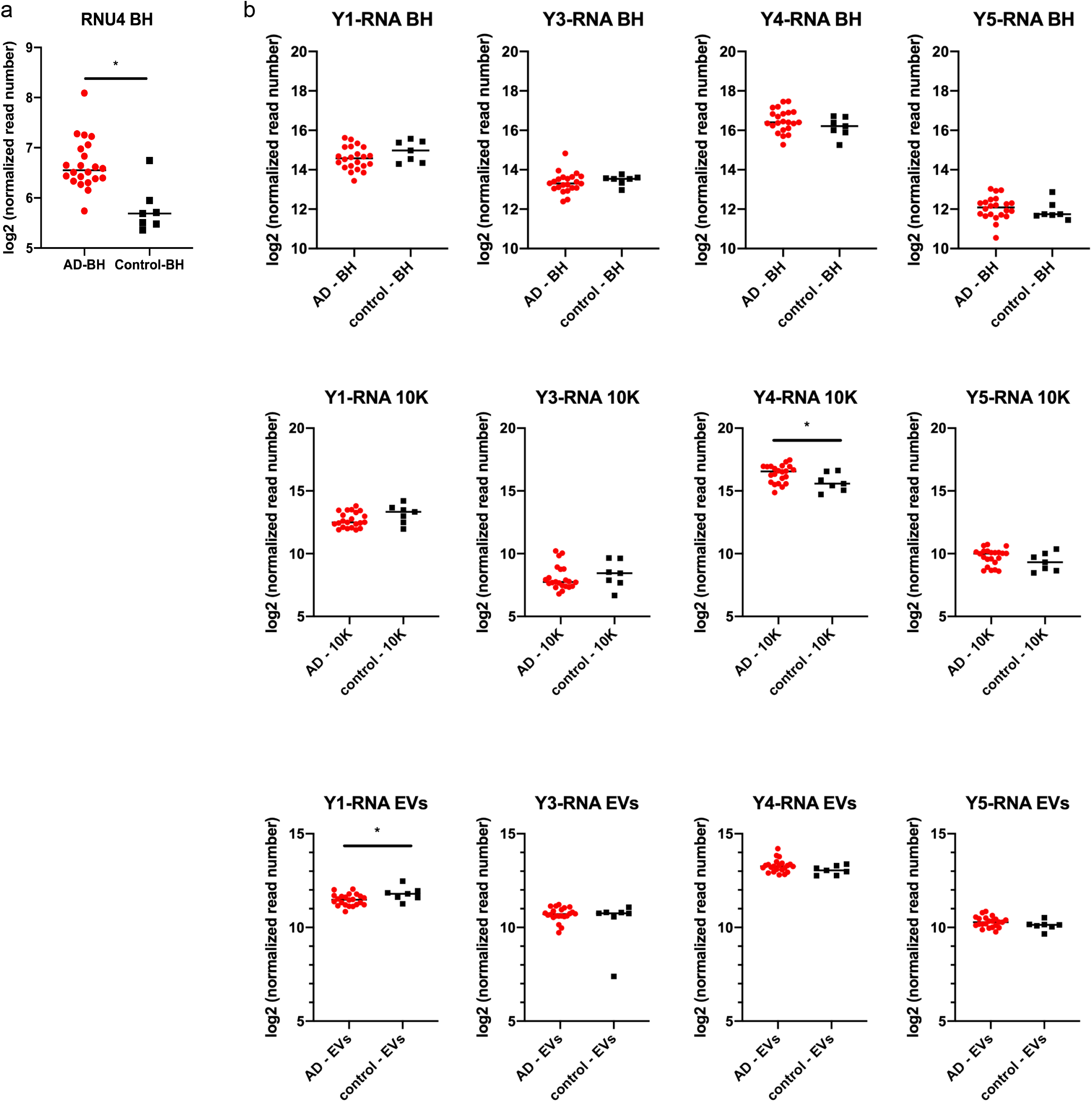
Differential expression of RNU and Y-RNAs related to AD pathology. (a) RNU4 expression in AD versus control in BH. Data are presented as mean +/- SD. (b) Y-RNAs differentially expressed in AD versus control in BH, 10K, and EVs. Data are presented as mean +/- SD. *p ≤ 0.05 as determined by DeSeq2 differential gene expression analysis based on the negative binomial distribution.

**Fig. S6.**
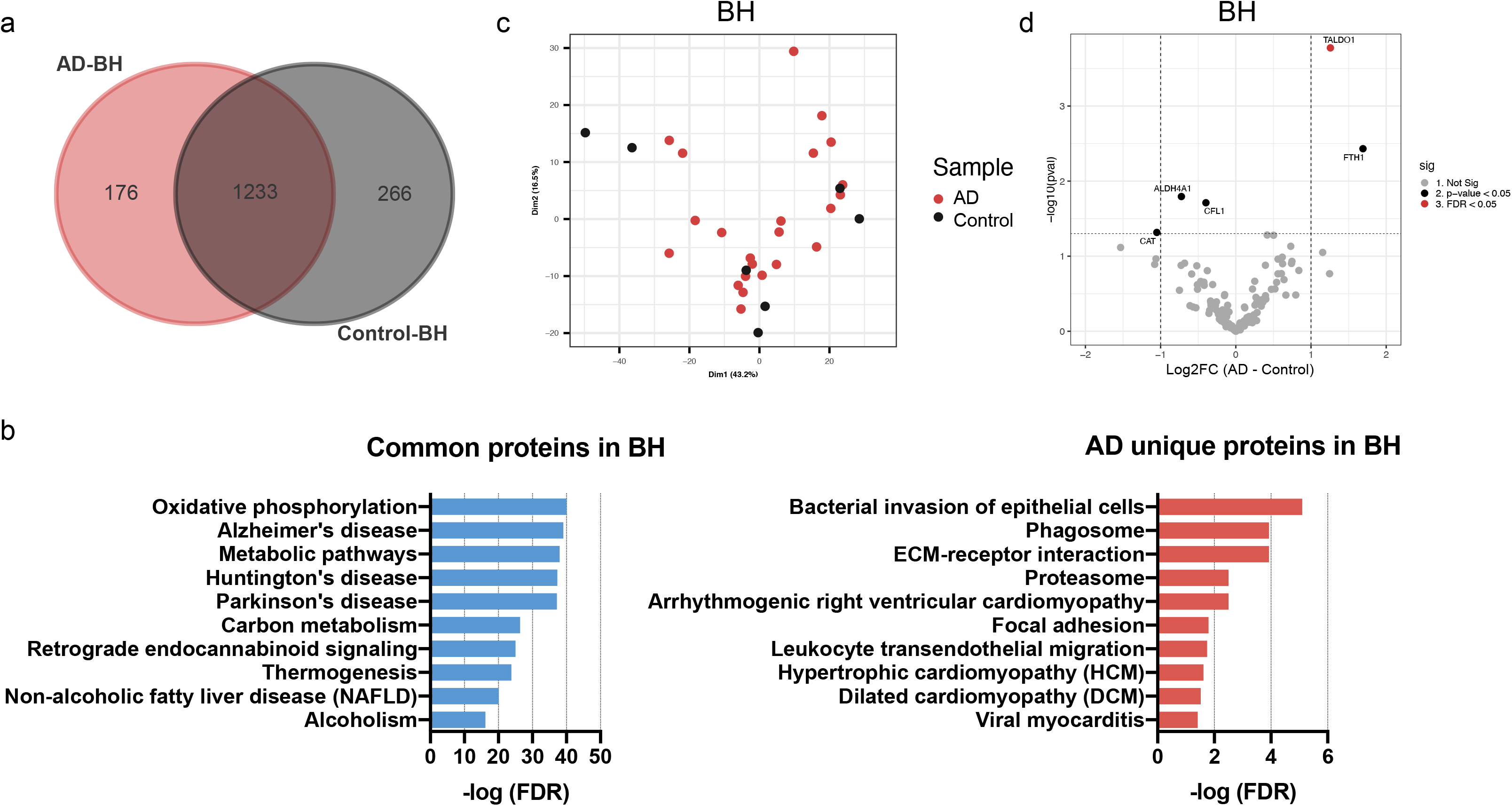
Proteins related to AD pathology in brain tissues. (a) Venn diagram of proteins identified in BH of AD and control. (b) The top 10 pathways ranked by FDR-corrected p value of BH proteins according to the Kyoto Encyclopedia of Genes and Genomes (KEGG). (c) Principal component analysis (PCA) based on proteome content of BH. (d) Volcano plots showing BH protein log2 fold changes (Log2FC) and p values (pval) for AD versus control. Thresholds for two-fold change and p value < 0.05 are indicated by dashed lines. Significant changes are indicated with different colors. Grey: non-significant (Not Sig), black: non-adjusted p value < 0.05, and red: FDR < 0.05.

**Fig. S7.**
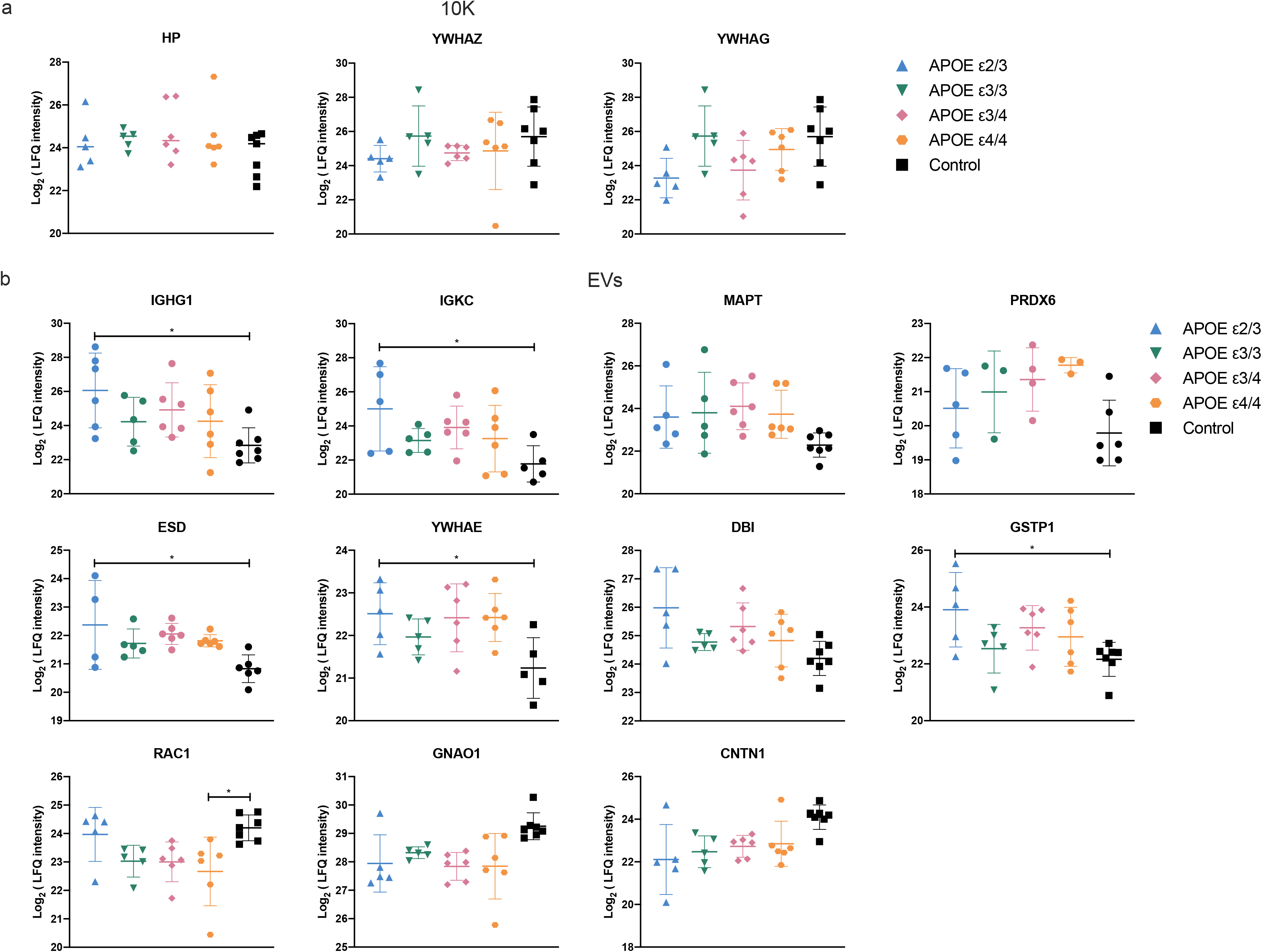
Protein related to APOE genotypes in brain tissues. (a) Venn diagram of proteins identified in BH from APOE ε4/4 and APOE ε2/3 carriers. (b) Volcano plots showing BH protein log2 fold changes (Log2FC) and p-value (pval) for APOE ε4/4 and APOE ε2/3 carriers. Thresholds for two-fold change and p value < 0.05 are indicated by dashed lines. Significant changes are indicated with different colors. Grey: non-significant (Not Sig), black: non-adjusted p value < 0.05, and red: FDR < 0.05. (c) The pathways significant for APOE ε4/4 and APOE ε2/3 unique proteins in BH according to the Kyoto Encyclopedia of Genes and Genomes (KEGG).

**Fig. S8.**
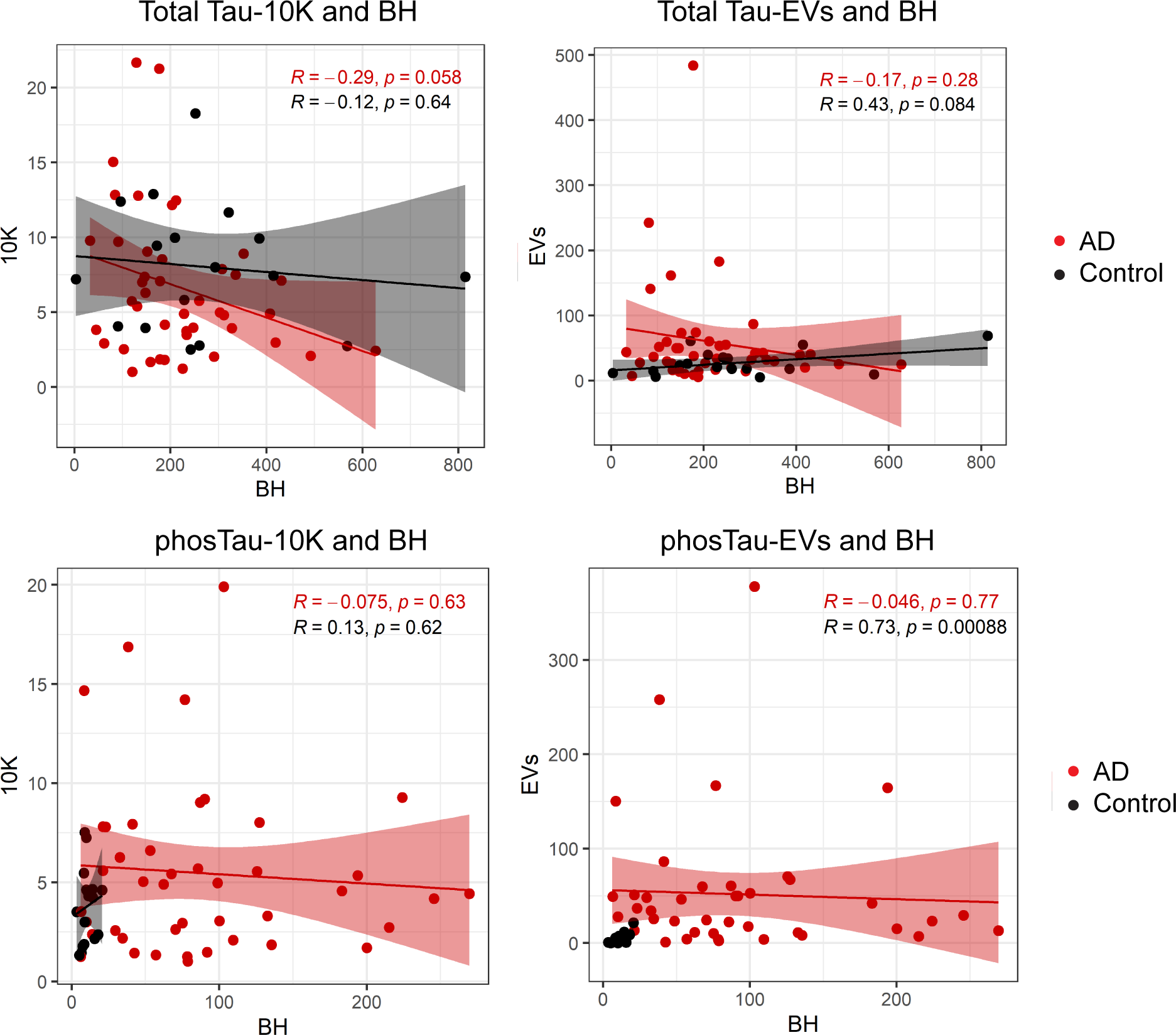
Correlations of Tau and phosphorylated Tau protein levels between BH with 10K (left), and BH with EVs (right) in AD and controls. Linear regression lines are shown in red and black for AD and control groups, respectively. The grey area depicts 95% confidence intervals. Pearson correlation coefficient (R) and significance (p) are shown based on n = 61 AD and control samples.

**Fig. S9.**
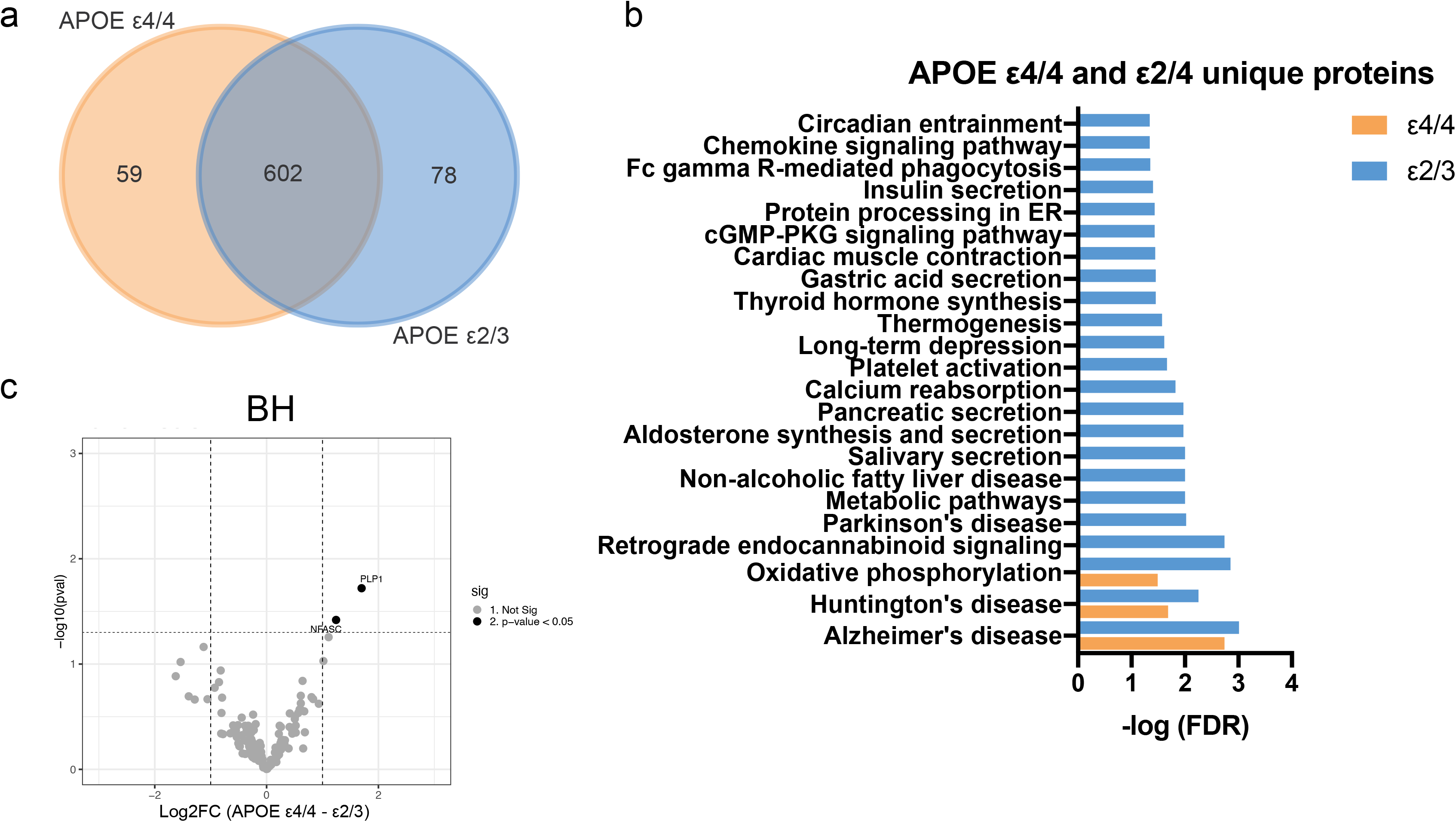
Protein related to APOE genotypes in brain tissues. (a) Venn diagram of proteins identified in BH from APOE ε4/4 and APOE ε2/3 carriers. (b) Volcano plots showing BH protein log2 fold changes (Log2FC) and p-value (pval) for APOE ε4/4 and APOE ε2/3 carriers. Thresholds for two-fold change and p value < 0.05 are indicated by dashed lines. Significant changes are indicated with different colors. Grey: non-significant (Not Sig), black: non-adjusted p value <0.05, and red: FDR < 0.05. (c) The pathways significant for APOE ε4/4 and APOE ε2/3 unique proteins in BH according to the Kyoto Encyclopedia of Genes and Genomes (KEGG).

**Fig. S10.**
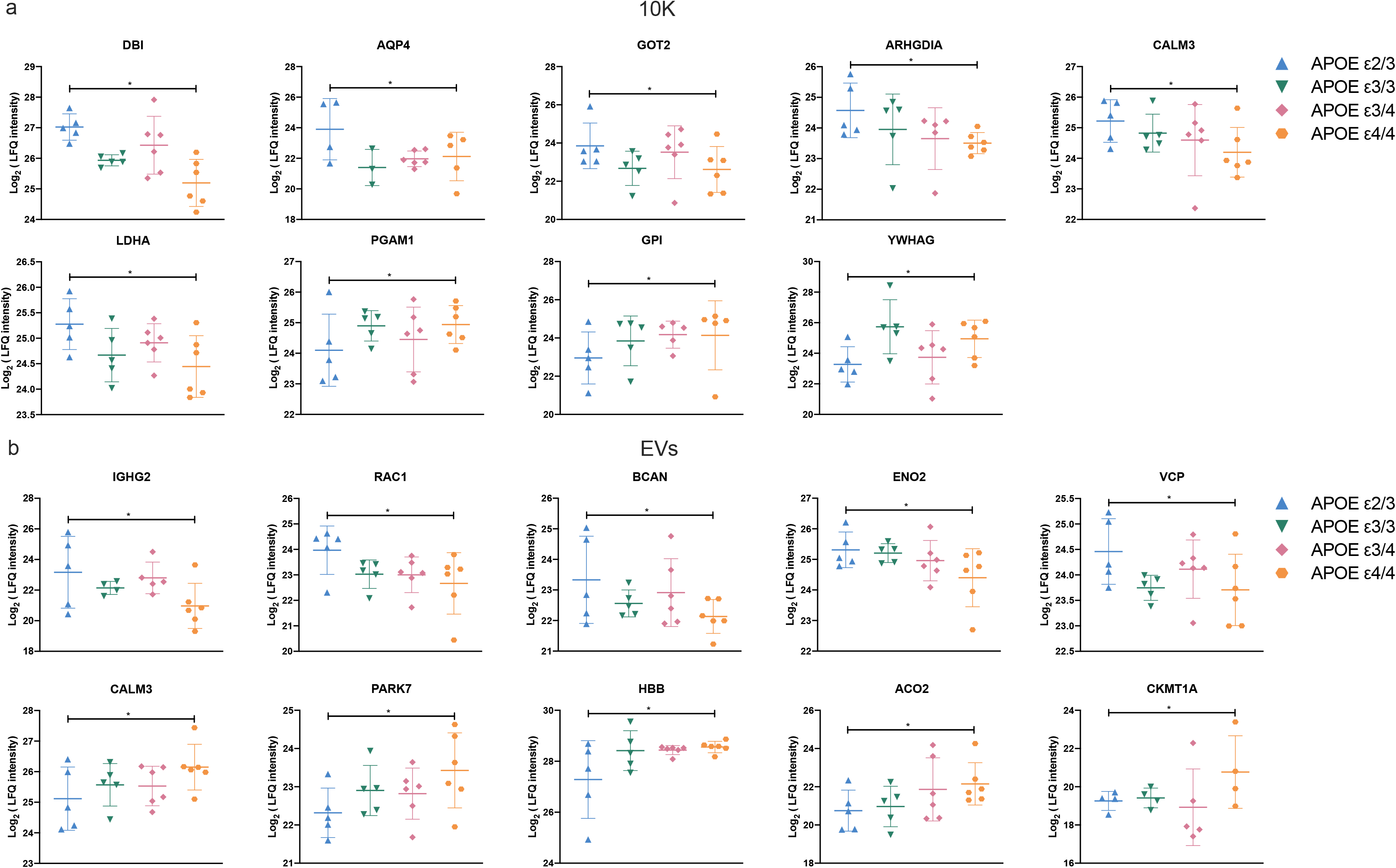
Expression level of proteins differentially expressed in APOE ε4/4 versus APOE ε2/3 for 10K (a) and EVs (b). Data are presented as mean +/- SD.

**Supplemental Table 1.** Donors: Cohort 1.

| Diagnosis | APOE genotype | Age | Sex (M<sup>a</sup>/F<sup>b</sup>) | RACE (W<sup>c</sup>/B<sup>d</sup>) | Braak Stage | CERAD stage | PMI<sup>e</sup> (h) |
| --- | --- | --- | --- | --- | --- | --- | --- |
| AD | E2/3 | 60s | M | W | 6 | C | 4 |
| AD | E2/3 | 60s | F | W | 6 | C | 11 |
| AD | E2/3 | 60s | M | W | 6 | C | 11 |
| AD | E2/3 | 80s | M | W | 5 | C | 5 |
| AD | E2/3 | 60s | M | W | 6 | C | 4 |
| AD | E3/3 | 80s | M | W | 6 | C | 7 |
| AD | E3/3 | 100s | F | W | 6 | B | 14 |
| AD | E3/3 | 80s | F | W | 6 | C | 8 |
| AD | E3/3 | 60s | M | W | 6 | C | 18 |
| AD | E3/3 | 80s | F | W | 6 | C | 17 |
| AD | E3/4 | 50s | F | W | 6 | C | 19 |
| AD | E3/4 | 90s | F | W | 6 | C | 21 |
| AD | E3/4 | 80s | F | W | 6 | C | 5 |
| AD | E3/4 | 80s | M | W | 6 | C | 9.5 |
| AD | E3/4 | 60s | F | W | 6 | C | 5.5 |
| AD | E3/4 | 80s | M | W | 6 | C | 18 |
| AD | E4/4 | 60s | M | W | / | / | 20 |
| AD | E4/4 | 90s | F | W | / | / | 6 |
| AD | E4/4 | 70s | F | W | / | / | 6 |
| AD | E4/4 | 90s | F | W | 6 | C | 6 |
| AD | E4/4 | 60s | F | W | / | / | 12 |
| AD | E4/4 | 70s | F | W | 6 | C | 13 |
| AD | / | 70s | M | W | 5 | C | 9 |
| Control | / | 80s | M | B | / | / | 10 |
| Control | / | 80s | M | W | / | / | 6 |
| Control | / | 50s | F | W | / | / | 6 |
| Control | E3/3 | 70s | M | W | 2 | / | 4 |
| Control | / | 90s | F | W | 1 | / | 8 |
| Control | / | 80s | F | W | 2 | / | 7 |
| Control | / | 90s | M | W | 2 | / | 15 |

<sup>a</sup> M = male; <sup>b</sup> F = female; <sup>c</sup> W = White; <sup>d</sup> B = Black or African American; <sup>e</sup> post-mortem interval

**Supplemental Table 2.**
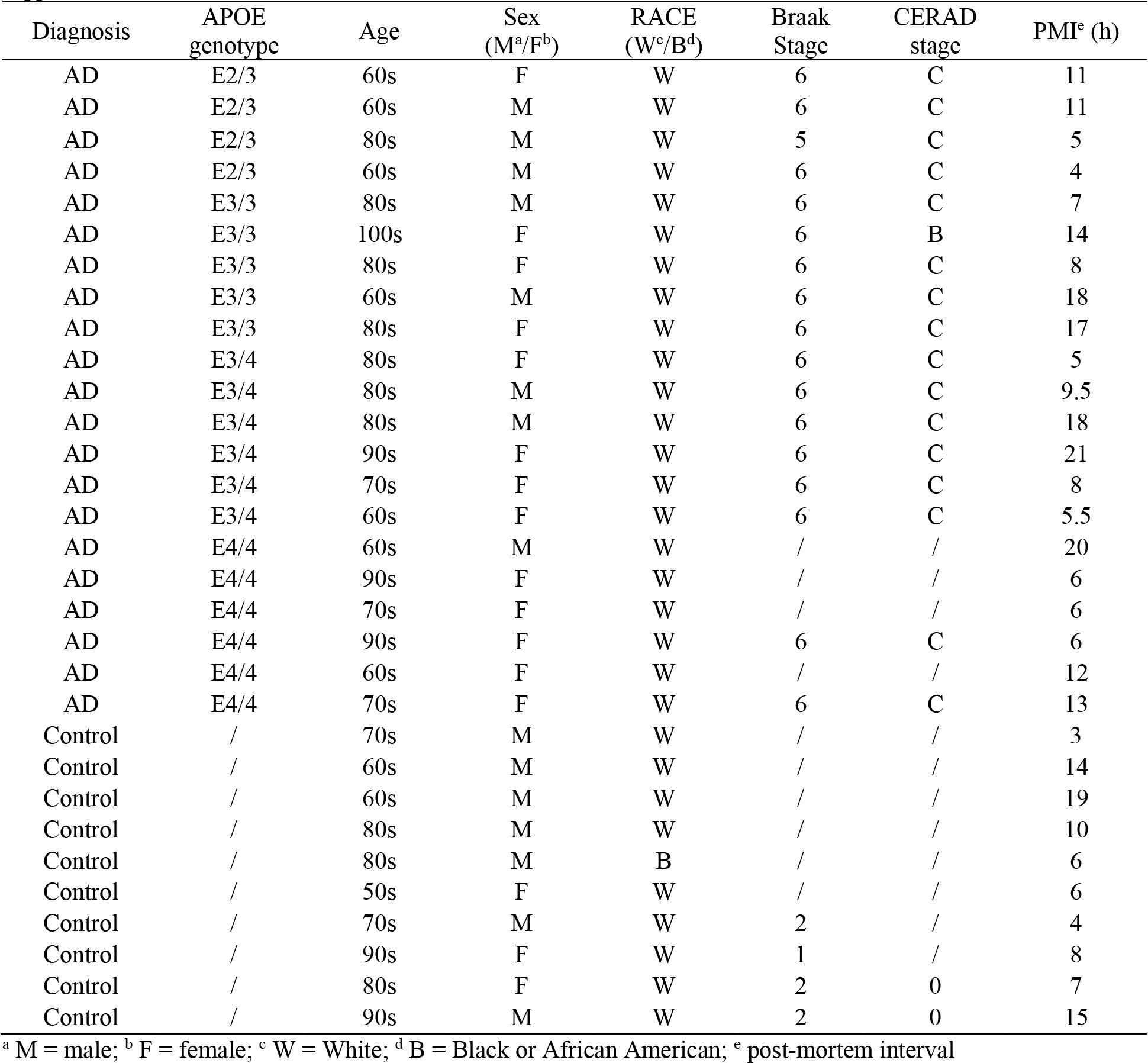
Donors: Cohort 2.

| Diagnosis | APOE genotype | Age | Sex (M<sup>a</sup>/F<sup>b</sup>) | RACE (W<sup>c</sup>/B<sup>d</sup>) | Braak Stage | CERAD stage | PMI<sup>e</sup> (h) |
| --- | --- | --- | --- | --- | --- | --- | --- |
| AD | E2/3 | 60s | F | W | 6 | C | 11 |
| AD | E2/3 | 60s | M | W | 6 | C | 11 |
| AD | E2/3 | 80s | M | W | 5 | C | 5 |
| AD | E2/3 | 60s | M | W | 6 | C | 4 |
| AD | E3/3 | 80s | M | W | 6 | C | 7 |
| AD | E3/3 | 100s | F | W | 6 | B | 14 |
| AD | E3/3 | 80s | F | W | 6 | C | 8 |
| AD | E3/3 | 60s | M | W | 6 | C | 18 |
| AD | E3/3 | 80s | F | W | 6 | C | 17 |
| AD | E3/4 | 80s | F | W | 6 | C | 5 |
| AD | E3/4 | 80s | M | W | 6 | C | 9.5 |
| AD | E3/4 | 80s | M | W | 6 | C | 18 |
| AD | E3/4 | 90s | F | W | 6 | C | 21 |
| AD | E3/4 | 70s | F | W | 6 | C | 8 |
| AD | E3/4 | 60s | F | W | 6 | C | 5.5 |
| AD | E4/4 | 60s | M | W | / | / | 20 |
| AD | E4/4 | 90s | F | W | / | / | 6 |
| AD | E4/4 | 70s | F | W | / | / | 6 |
| AD | E4/4 | 90s | F | W | 6 | C | 6 |
| AD | E4/4 | 60s | F | W | / | / | 12 |
| AD | E4/4 | 70s | F | W | 6 | C | 13 |
| Control | / | 70s | M | W | / | / | 3 |
| Control | / | 60s | M | W | / | / | 14 |
| Control | / | 60s | M | W | / | / | 19 |
| Control | / | 80s | M | W | / | / | 10 |
| Control | / | 80s | M | B | / | / | 6 |
| Control | / | 50s | F | W | / | / | 6 |
| Control | / | 70s | M | W | 2 | / | 4 |
| Control | / | 90s | F | W | 1 | / | 8 |
| Control | / | 80s | F | W | 2 | 0 | 7 |
| Control | / | 90s | M | W | 2 | 0 | 15 |

<sup>a</sup> M = male; <sup>b</sup> F = female; <sup>c</sup> W = White; <sup>d</sup> B = Black or African American; <sup>e</sup> post-mortem interval

**Supplemental Table 3.**
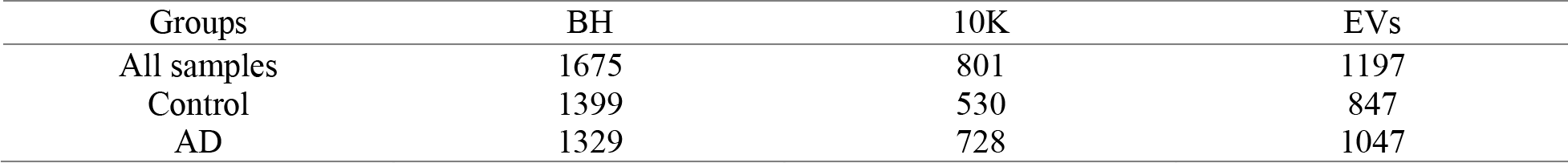
Protein number identified in samples.

| Groups | BH | 10K | EVs |
| --- | --- | --- | --- |
| All samples | 1675 | 801 | 1197 |
| Control | 1399 | 530 | 847 |
| AD | 1329 | 728 | 1047 |

